# Prevalence and patient characteristics associated with cardiovascular disease risk factor screening in UK primary care for people with severe mental illness: An electronic healthcare record study

**DOI:** 10.1101/2024.10.09.24315173

**Authors:** Launders N, Jackson CA, Hayes JF, John A, Stewart R, Iveson MH, Bramon E, Guthrie B, Mercer SW, Osborn DPJ

**Affiliations:** Division of Psychiatry, University College London, London, United Kingdom; Usher Institute, The University of Edinburgh, Edinburgh, United Kingdom; Camden and Islington NHS Foundation Trust, London, United Kingdom; Swansea University Medical School, Swansea University, Swansea, United Kingdom; Institute of Psychiatry, Psychology and Neuroscience, Kings College London, London, United Kingdom; South London and Maudsley NHS Foundation Trust, London, UK; Division of Psychiatry, Centre for Clinical Brain Sciences, University of Edinburgh, Edinburgh, United Kingdom

## Abstract

**Background:** People with severe mental illness (SMI) are at increased risk of cardiovascular disease (CVD), and initiatives for CVD risk factor screening in the UK have not reduced disparities.

**Objectives:** To describe the annual screening prevalence for CVD risk factors in people with SMI from April 2000 to March 2018, and to identify factors associated with receiving no screening and regular screening.

**Methods:** We identified adults with a diagnosis of SMI (schizophrenia, bipolar disorder or ‘other psychosis’) from UK primary care records in Clinical Practice Research Datalink (CPRD). We calculated the annual prevalence of screening for blood pressure, cholesterol, glucose, body mass index, alcohol consumption and smoking status, using multinomial logistic regression to identify factors associated with receiving no screening and complete screening.

**Results:** Of 216,136 patients with SMI, 55% received screening for all six CVD risk factors at least once follow-up and 35% received all six within a one-month period. Changes in screening prevalence coincided with changes in incentivisation of screening. In 2014-2018, men, people with a diagnosis of ‘other psychoses’, or with missing ethnicity were more likely to have received no screening.

**Conclusions:** The low proportion of people with SMI receiving regular comprehensive CVD risk factor screening is concerning. Screening needs to be embedded as part of broad physical health checks to ensure the health needs of people with SMI are being met. If we are to improve cardiovascular health, interventions are needed where risk of receiving no screening or not receiving regular screening is highest.

**What is already known on this topic**

- The prevalence of CVD risk factor screening in primary care increased with the introduction of an incentivisation scheme and when incentivisation for screening for specific CVD risk factors is removed the screening decreases.

**What does this study add?**

- Few patients received regular screening for all six CVD risk factors considered, and both screening prevalence and risk of receiving no screening varied depending on patient characteristics.
- Only 35% of patients ever received screening for all six CVD risk factors considered in a one-month period, suggesting that screening is not often being done as part of a physical health check.

**How this study might affect research, practice or policy**

- Clinicians should be aware that some sub-groups of patients with SMI are less likely to receive screening, and the importance of providing screening as part of a regular, comprehensive physical health check.

## Background

People with severe mental illnesses (SMI), such as schizophrenia, bipolar disorder or other psychotic illnesses, are at increased risk of many physical health conditions^1^ ^2^. People with SMI have 1.5 to 2.5 times the risk of cardiovascular disease (CVD) compared to the general population and an increased risk of death from CVD^3–7^. They also have a higher prevalence of CVD risk factors, such as smoking, obesity and diabetes^1^ ^8–10^, which is compounded by cardiometabolic side effects of antipsychotic medication^11^, and sociodemographic risk factors^12^ ^13^.

In recognition of physical health disparities in people with SMI, financial incentivisation of physical health checks for people with SMI in primary care was introduced in the UK in 2004 through the Qualities and Outcomes Framework (QOF)^14^. QOF initially incentivised a review of physical health, with incentivisation of screening for individual CVD risk factors introduced in 2011. While blood pressure and alcohol consumption screening have been consistently incentivised since 2011 and smoking status has been incentivised since 2008, cholesterol, glucose and BMI screening have been less consistently incentivised (see Table S1 for QOF changes related to SMI). Additionally, from 2014 the measures incentivised differed across the constituent countries of the UK, and Scotland and Wales abolished QOF in 2016 and 2019 respectively.

Several studies have shown an increase in recording of CVD risk factors following the introduction of QOF in people with SMI^15–18^, and a recent cohort study found that in England, removal of cholesterol and BMI as incentivised indicators resulted in a decrease in recording of these risk factors compared to blood pressure recording^19^. However, there is a lack of evidence regarding long term trends in screening prevalence for the six CVD risk factors currently included in the NHS England Physical Health Check for SMI, and patient characteristics associated with receiving screening. In order to identify unmet need and improve the health of people with SMI, it is important to understand whether incentivisation drives increases in screening in all patients with SMI, and to identify which individuals may be at risk of not being screened.

Objective: To investigate the long-term trends and patient characteristics associated with receipt of comprehensive CVD risk factor screening: blood pressure, cholesterol, glucose screening, BMI measurement, alcohol consumption and smoking status.

## Methods

### Study design

We used Clinical Practice Research Datalink (CPRD) GOLD and Aurum databases to patients with SMI. These databases contain deidentified healthcare records for patients registered with primary care practices in the UK and are broadly representative of the UK population^20^ ^21^. Our protocol was pre-registered (https://osf.io/czetb/). We investigated annual screening prevalence of six CVD risk factors at the population level. We then investigated screening patterns at an individual level and patient characteristics associated with receipt of screening.

### Population

We identified patients aged over 18 with a diagnosis of SMI (defined as schizophrenia, bipolar disorder or other non-organic psychosis) recorded in primary care records. Entry to the cohort was the latest of SMI diagnosis date, registration at primary care practice, or 1 April 2000. Exit was the earliest of death, leaving the primary care practice, 100^th^ birthday, or 31 March 2018. Patients did not re-enter the cohort following exit.

Patients were required to have at least one year of follow-up following entry into the cohort to allow time for screening to be recorded. In line with QOF reporting rules patients had to be registered with their primary care practice for the last three months of a financial year in order to be eligible for screening in that year. In the analysis of factors associated with CVD risk factor screening, we stratified the follow-up time into three periods; April 2004- March 2011; April 2011 – March 2014; April 2014 – March 2018, based on major changes to the QOF incentivisation programme (Table S1). Patients were included in each period if they were eligible for screening for at least two financial years of that period.

### Covariates

We defined the following covariates *a priori*:

We defined sex, primary care practice and country of primary care practice as recorded in CPRD, and prescription of antipsychotics or mood stabilisers (lithium, sodium valproate or lamotrigine) based on recorded prescriptions issued in primary care.

We defined age as under 40 years or 40 years and older based on year of birth, chosen because cholesterol and blood glucose screening incentivisation was limited to those 40 years and over^22^. In multinomial logistic regression models, we included age at start of follow-up as a continuous variable and reported effect size as odds ratios (OR) per 10-year increase in age.

We defined ethnicity as recorded in primary care records and grouped as per the UK 2011 Census^23^. We defined specific SMI diagnosis was defined as the most recently recorded of schizophrenia, bipolar disorder or other non-organic psychotic illness.

We defined presence on other QOF registers which incentivise CVD risk factor screening as having a Read code used in the 2017-2018 QOF incentivisation for any of: atrial fibrillation, coronary heart disease, hypertension, peripheral artery disease, stroke or diabetes. Exception reporting was defined as ever having a Read code that would remove a patient from the incentivisation denominator for the mental health domain (i.e. deemed unsuitable, withdrew consent, or didn’t respond to invitations for screening).

### Outcomes

We investigated screening of six individual CVD risk factors (cholesterol, blood glucose, blood pressure, BMI, smoking status and alcohol consumption screening), and a composite outcome of all six. For glucose, we included codes for blood glucose or HbA1C tests or values, but excluded urine testing. For cholesterol, we included any screening code or value. For blood pressure, we included screening codes or values for either diastolic or systolic blood pressure. For BMI we included BMI values, BMI calculated from height and weight, and screening codes. For smoking and alcohol status we included any screening code. Further details on the prevalence of CVD risk factors in this cohort is available on the DATAMIND website (https://datamind.org.uk/data/harmonised-data/smi-cohorts/) and code lists used to define the population, covariates and outcomes are available in the HDR UK phenotype library (Table S2).

In the individual-level analysis of factors associated with CVD risk factor screening, in each time period a patient was considered to have ‘always complete screening’ if they received screening for all six CVD risk in each financial year in the time period that they were active. Irregular screening was defined as any frequency between ‘always complete screening’ and receiving no screening.

### Statistical analysis

We calculated the annual prevalence of recorded screening of each of the six CVD risk factors, stratifying by the aforementioned covariates. We calculated the proportion of patients receiving all six CVD risk factors ever during follow-up, ever within a one-month period, and for each financial year.

We conducted multinomial logistic regression analyses to assess patient factors associated with receiving ‘always complete’ CVD risk factor screening and receiving no screening in each of the three time periods, compared to receiving irregular screening.

We mutually adjusted for all covariates in the models, with the exception of primary care practice which was used as a clustering term in the calculation of sandwich standard errors. We additionally adjusted for the time since SMI diagnosis, time since primary care practice registration, year of end of follow-up and total follow-up time. Analysis was performed in R and R Studio and reported in line with the RECORD checklist^24^.

### Missing data

Missing ethnicity was included as a separate category as those with missing ethnicity are different to those with a recorded ethnicity with respect to healthcare access and engagement. For all diagnostic and screening variables, we deemed absence of a code to indicate an absence of diagnosis or screening. We excluded 122 patients who were missing geographical data from the analysis.

### Sensitivity analyses

In *a priori* sensitivity analysis we limited the population to patients resident in England with available deprivation data (English Index of Multiple Deprivation (IMD) quintiles). In *post-hoc* analysis we limited the model to patients who were active for the whole of the follow-up period due to the strong effect of follow-up time on the completeness of screening.

### PPI involvement

Lived experience advisors from the DATAMIND Super Research Advisory Group (https://datamind.org.uk/patients-and-public/the-super-research-advisory-group/) and UCL Mental Health Data Science PPIE group commented on the protocol and provided input into the interpretation of results.

### Findings

We identified 312,471 patients with a diagnostic code for SMI at any time, of whom 216,136 had a diagnosis of SMI before the end of follow-up, were over the age of 18 years, with at least one year of registration and without missing geographic data (Figure S1). Patients had a median of 4.85 (IQR: 2.43, 9.72) years of follow-up (Table 1).

**Table 1:** Characteristics of the cohort of patients with severe mental illness in CPRD, n=216,136.

| Characteristic |  | n (%) / median [IQR] |
| --- | --- | --- |
| **Age at SMI diagnosis (median [IQR])** |  | 35 [26, 48] |
| **Age at start of follow-up (median [IQR])** |  | 44 [33, 59] |
| **Age at end of follow-up (median [IQR])** |  | 52 [39, 68] |
| **Follow-up time<sup>a</sup> (median [IQR])** |  | 4.85 [2.43, 9.72] |
| **Sex, n (%)** | Male | 111,655 (51.7) |
|  | Female | 104,481 (48.3) |
| **Ethnicity, n (%)** | Asian | 7,679 (3.6) |
|  | Black | 9,979 (4.6) |
|  | Mixed | 2,733 (1.3) |
|  | Other | 4,199 (1.9) |
|  | White | 110,673 (51.2) |
|  | Missing | 80,873 (37.4) |
| **Country, n (%)** | England | 186,880 (86.5) |
|  | Northern Ireland | 3,405 (1.6) |
|  | Scotland | 14,010 (6.5) |
|  | Wales | 11,841 (5.5) |
| **Most recent SMI diagnosis, n (%)** | Schizophrenia | 73,753 (34.1) |
|  | Bipolar disorder | 68,921 (31.9) |
|  | Other psychoses | 73,462 (34.0) |
| **Ever exception reported, n (%)** |  | 59,736 (27.6) |
| **Ever on another CVD QOF register<sup>b</sup>, n (%)** |  | 64,295 (29.8) |
| **Ever prescribed antipsychotics, lithium, sodium valproate or lamotrigine, n (%)** |  | 173,669 (80.3) |
| **Died during follow-up<sup>a</sup>, n (%)** |  | 31,210 (14.4) |
| **Age at death (median [IQR])** |  | 75.49 [62.44, 84.74] |

SMI: Severe Mental Illness; CVD: cardiovascular disease; IQR: interquartile range; QOF: Quality and Outcomes Framework
a: Follow-up starts at the latest of 01/04/2000, primary care practice registration, SMI diagnosis or age 18 and ends at the earliest of death, leaving the primary care practice, age 100 or last data collection by CPRD.
b: Defined as presence on QOF register for atrial fibrillation, coronary heart disease, hypertension, peripheral artery disease, stroke or diabetes.

### Population-level analysis of cardiovascular disease risk factor screening

The prevalence of smoking and blood pressure screening increased steadily during the study period. In contrast, for alcohol, BMI, cholesterol and glucose screening the prevalence of screening increased sharply in 2011-2012 following the introduction of incentivisation of individual CVD risk factors. For BMI, cholesterol and glucose screening the prevalence decreased rapidly from 2013-2014 to 2014- 2015, coinciding with the withdrawal of financial incentives (Figure 1, Table S3).

**Figure 1:**
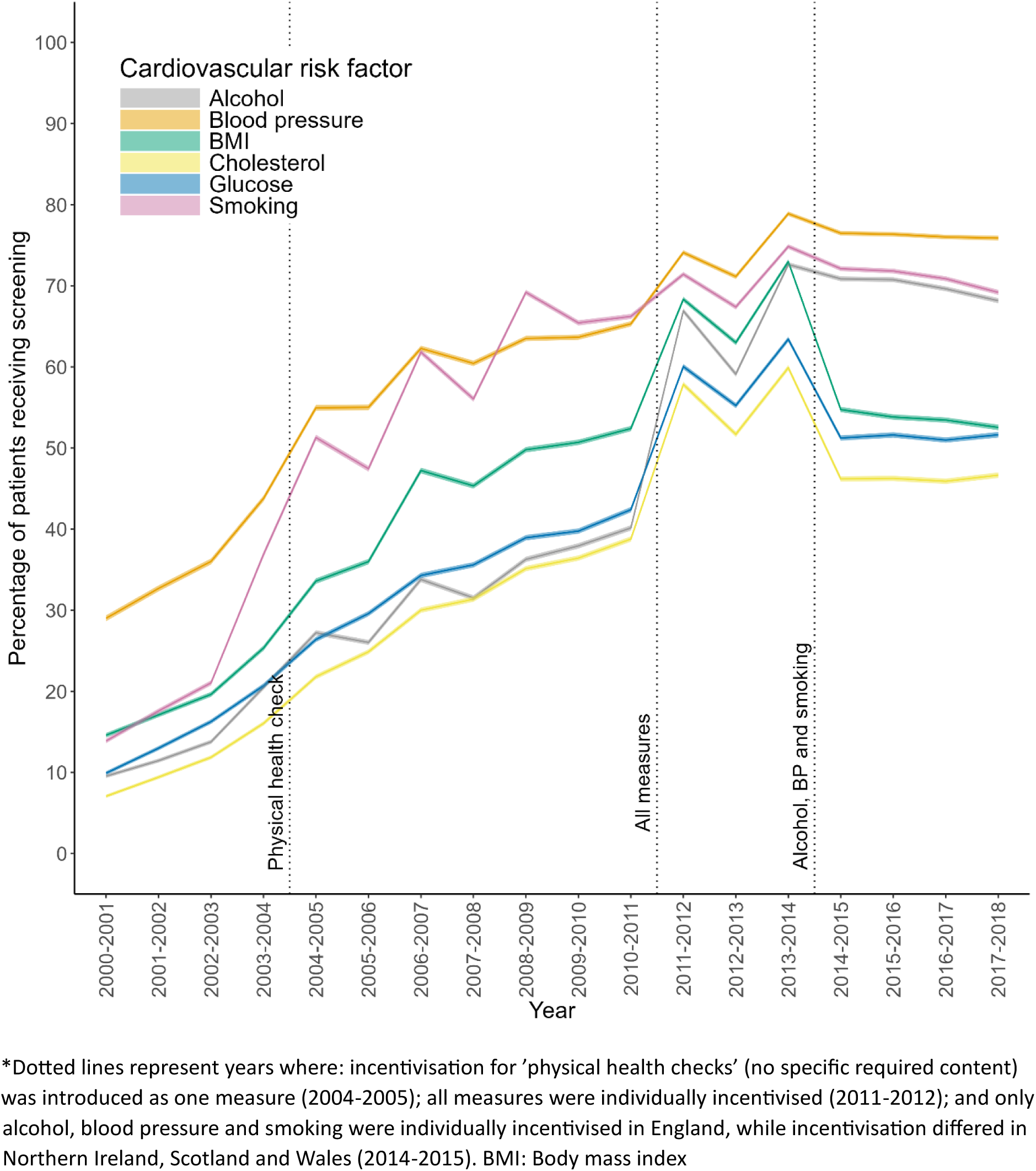
Prevalence of cardiovascular risk factor screening in people with severe mental illness, in the primary care setting in the UK by financial year

These broad patterns remained when stratified by most patient characteristics (Figures S2-S10). However, the increase in cholesterol and glucose screening in 2011-2013 was primarily observed in those aged 40 years or older (Figure S2), the population incentivised at the time. While the prevalence of screening increased for all countries from 2000 to 2014, from 2014 to 2018 (a period of diverging incentivisation across the four nations) the pattern was less consistent (Figure S3).

Screening for all CVD risk factors was lowest in those with a diagnosis of ‘other psychoses’ (Figure S4), men (Figure S5), those not on another QOF register (Figure S6) and those not on antipsychotics or mood stabilisers (Figure S7). Screening prevalence for smoking was highest in patients of White or Mixed ethnicity, and screening of other CVD risk factors was highest in patients of Asian ethnicity (Figure S7).

### Individual-level factors associated with receiving ‘always complete’ or no CVD risk factor screening

Almost all patients (93.9%) received screening for at least one of the six CVD risk factors at some point during follow-up. However, only half (54.8%) received screening for all six CVD risk factors at least once (Table 2), and this occurred within a one-month period at least once for only 34.8% of patients.

**Table 2:**
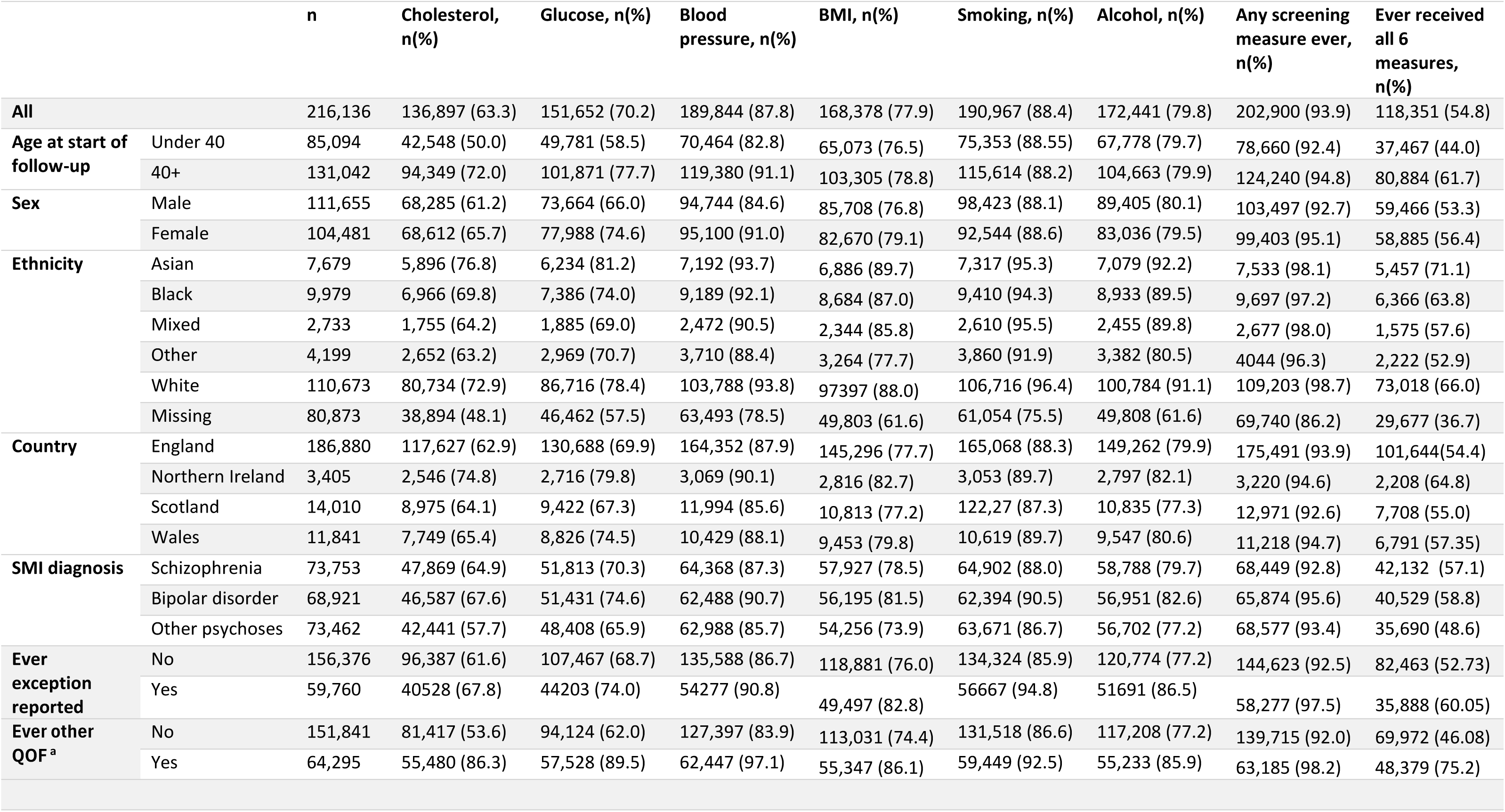
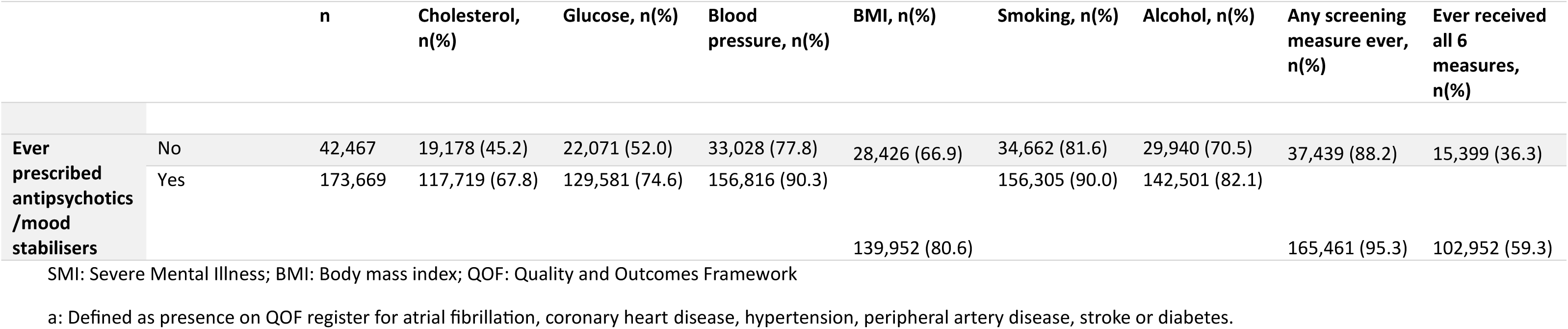
Proportion of patients with severe mental illness ever receiving screening for: each cardiovascular disease (CVD) risk factor; any of the six CVD risk factors; and all six CVD risk factors, stratified by covariates, 2000-2018.

In the period prior to all six CVD risk factors being incentivised (2004-2011) 1.7% of patients received ‘always complete’ screening (i.e. all six CVD risk factors each financial year that they were active between 2004 and 2011). This increased to 14.8% during the period of individual incentivisation (2011-2014) and decreased to 8.3% following that (2014-2018; Table 3).

**Table 3:** Unadjusted multinomial logistic regression for the odds ratio of always receiving complete screening^a^ or receiving no screening compared to irregular screening, among people with severe mental illness, during two time periods.

|  |  | April 2011 – March 2014 (n=85,274) |  | April 2014 – March 2018 (n=94,216) |  |
| --- | --- | --- | --- | --- | --- |
| Reference: Irregular screening |  | Complete<br>(n=12,616, 14.79%) | None (n=3,204,<br>3.76%) | Complete<br>(n=7,771, 8.25%) | None (n=2,092,<br>2.22%) |
|  |  | OR (95% CI) | OR (95% CI) | OR (95% CI) | OR (95% CI) |
| Age at start of follow-up | Per 10-year increase | **1.26 (1.24-1.27)** | 1.00 (0.95-1.06) | **1.27 (1.24-1.28)** | **0.82 (0.79-0.94)** |
| Sex (ref female) | Male | 1.03 (0.98-1.07) | **1.58 (1.46-1.72)** | **1.09 (1.04-1.15)** | **1.93 (1.75-2.12)** |
| Ethnicity (ref White) | Asian | **1.13 (1.00-1.28)** | 1.02 (0.81-1.30) | **1.54 (1.34-1.78)** | 0.82 (0.62-1.09) |
|  | Black | **0.81 (0.71-0.92)** | **1.60 (1.27-2.02)** | **1.18 (1.01-1.37)** | **1.35 (1.08-1.68)** |
|  | Mixed | **0.65 (0.53-0.78)** | 1.24 (0.85-1.80) | 0.83 (0.67-1.01) | 0.89 (0.58-1.36) |
|  | Other | **0.68 (0.57-0.81)** | **2.43 (1.88-3.13)** | **0.73 (0.58-0.93)** | **2.12 (1.61-2.79)** |
|  | Missing | **0.73 (0.66-0.80)** | **4.76 (3.92-5.78)** | **0.68 (0.61-0.75)** | **2.62 (2.33-2.95)** |
| Country (ref England) | Northern Ireland | 1.04 (0.69-1.57) | **0.55 (0.37-0.79)** | **0.58 (0.35-0.94)** | 1.31 (0.90-1.90) |
|  | Scotland | **1.36 (1.17-1.59)** | 0.85 (0.68-1.07) | 1.06 (0.86-1.30) | **3.02 (2.59-3.52)** |
|  | Wales | **0.66 (0.55-0.79)** | 0.80 (0.61-1.58) | **0.52 (0.43-0.65)** | **1.95 (1.57-2.42)** |
| SMI diagnosis (ref bipolar disorder) | Schizophrenia | **1.26 (1.20-1.32)** | **1.31 (1.19-1.45)** | **1.38 (1.30-1.48)** | **1.17 (1.04-1.32)** |
|  | Other psychoses | **0.83 (0.78-0.87)** | **1.44 (1.27-1.63)** | **0.87 (0.82-0.93)** | **1.82 (1.63-2.03)** |
| In period variables<sup>d</sup> | Exception reported<sup>b</sup> | **0.37 (0.35-0.39)** | **1.46 (1.19-1.80)** | **0.45 (0.42-0.48)** | **2.14 (1.92-2.38)** |
|  | Other QOF register<sup>c</sup> | **2.30 (2.17-2.42)** | **0.28 (0.25-0.31)** | **3.71 (3.39-4.07)** | **0.28 (0.52-0.30)** |
|  | On antipsychotics/mood stabilisers | **2.05 (1.93-2.18)** | **0.14 (0.13-0.16)** | **2.14 (1.97-2.32)** | **0.21 (0.19-0.23)** |
|  | Years since diagnosis | 1.00 (1.00-1.00) | 1.00 (1.00-1.00) | 1.00 (1.00-1.00) | 1.00 (1.00-1.00) |
|  | Years since registration | 1.00 (1.00-1.00) | 1.00 (1.00-1.00) | 1.00 (1.00-1.00) | 1.00 (1.00-1.00) |
|  | Years of follow-up | 1.00 (1.00-1.00) | 0.998 (0.998-0.999) | 1.00 (1.00-1.00) | 0.999 (0.999-1.00) |
| Year of end of record in period (ref final year)<sup>e</sup> | Year 2 | **0.00 (00.00-0.00)** | **0.00 (00.00-0.00)** | 0.43 (0.57-3.15) | **5.07 (1.52-16.87)** |
|  | Year 3 | NA | NA | **1.66 (1.51-1.83)** | **2.50 (2.19-2.86)** |

OR: Odds ratio; 95% CI: 95% confidence interval; ref: Reference category; SMI: Severe Mental Illness; QOF: Quality and Outcomes Framework
a: Complete screening was defined as screening of all six cardiovascular risk factors for each year that the patient is active
b: Exception reported from mental health measures
c: Defined as presence on QOF register for atrial fibrillation, coronary heart disease, hypertension, peripheral artery disease, stroke or diabetes. d: In period variables measured cross-sectionally, up to the end of the period of interest e: Defined as the year a patient ends follow-up. For the 2014-2018 cohort, year two is 2015-2016, year 3 is 2016-2017 and the final year is 2017-2018. For the 2011-2014 cohort year 2 is 2012-2013 and the final year is 2013-2014. Note: Patients could be present in both time periods. The total number of unique patients is 119,976

The odds of receiving no screening in the 2014-2018 and 2011-2014 periods were higher for men, patients of ‘other’ or missing ethnicity (compared to White ethnicity) and those who had been exception reported from QOF, even after mutual adjustment for covariates (Tables 3 & 4). Conversely, those on other QOF registers which incentivise screening or who had been prescribed antipsychotics or mood stabilisers were less likely to receive no screening in each time period (Tables 3 & 4). In adjusted analyses, compared to patients in England, patients resident in Scotland or Wales were less likely to receive no screening in the 2011-2014 period, but more likely to receive no screening in the 2014-2018 period when both countries reduced incentives in varying ways (Table 4, Table S1).

**Table 4:** Adjusted multinomial logistic regression^a^ for the odds ratio of always receiving complete screening^a^ or receiving no screening compared to irregular screening, among people with severe mental illness, during two time periods.

|  |  | April 2011 – March 2014 (n=85,274) |  | April 2014 – March 2018 (n=94,216) |  |
| --- | --- | --- | --- | --- | --- |
| Reference: Irregular screening |  | Complete (n=12,616, 14.79%) | None (n=3,204, 3.76%) | Complete (n=7,771, 8.25%) | None (n=2,092, 2.22%) |
| Age at start of follow-up | Per 10-year increase | **1.18 (1.17-1.21)** | 1.01 (0.96-1.05) | **1.15 (1.14-1.17)** | **0.86 (0.83-0.90)** |
| Sex (ref female) | Male | **1.24 (1.19-1.30)** | **1.31 (1.21-1.43)** | **1.33 (1.26-1.41)** | **1.41 (1.27-1.56)** |
| Ethnicity (ref White) | Asian | **1.24 (1.09-1.41)** | 1.00 (0.78-1.29) | **1.75 (1.51-2.04)** | 0.88 (0.66-1.17) |
|  | Black | 0.93 (0.82-1.06) | **1.31 (1.03-1.67)** | **1.39 (1.19-1.62)** | 1.23 (0.98-1.55) |
|  | Mixed | **0.81 (0.67-0.97)** | 1.02 (0.69-1.50) | 1.04 (0.85-1.28) | 0.83 (0.54-1.28) |
|  | Other | **0.77 (0.65-0.92)** | **2.56 (1.98-3.31)** | 0.83 (0.67-1.03) | **1.66 (1.29-2.15)** |
|  | Missing | **0.69 (0.63-0.76)** | **5.26 (4.34-6.38)** | **0.70 (0.63-0.78)** | **2.15 (1.89-2.44)** |
| Country (ref England) | Northern Ireland | 1.14 (0.75-1.73) | **0.33 (0.22-0.48)** | **0.60 (0.38-0.97)** | 1.17 (0.78-1.74) |
|  | Scotland | **1.56 (1.32-1.86)** | **0.67 (0.53-0.84)** | 1.15 (0.93-1.44) | **2.88 (2.43-3.41)** |
|  | Wales | **0.81 (0.67-0.97)** | **0.48 (0.36-0.64)** | **0.65 (0.54-0.79)** | **1.51 (1.21-1.88)** |
| SMI diagnosis (ref bipolar disorder) | Schizophrenia | **1.27 (1.20-1.34)** | 0.99 (0.89-1.10) | **1.30 (1.22-1.39)** | 0.90 (0.79-1.03) |
|  | Other psychoses | **0.92 (0.88-0.97)** | 1.05 (0.94-1.17) | 0.96 (0.90-1.02) | **1.22 (1.09-1.37)** |
| In period variables<sup>d</sup> | Exception reported<sup>b</sup> | **0.38 (0.36-0.41)** | **1.26 (1.08-1.47)** | **0.47 (0.44-0.51)** | **1.69 (1.51-1.89)** |
|  | Other QOF register<sup>c</sup> | **1.93 (1.82-2.04)** | **0.31 (0.28-0.35)** | **3.21 (2.93-3.53)** | **0.35 (0.31-0.39)** |
|  | On antipsychotics/mood stabilisers | **1.89 (1.77-2.01)** | **0.17 (0.15-0.19)** | **1.95 (1.80-2.11)** | **0.24 (0.21-0.26)** |
|  | Years since diagnosis | 1.00 (1.00-1.00) | 1.00 (1.00-1.00) | 1.00 (1.00-1.00) | 1.00 (1.00-1.00) |
|  | Years since registration | 1.00 (1.00-1.00) | 1.00 (1.00-1.00) | 1.00 (1.00-1.00) | 1.00 (1.00-1.00) |
|  | Years of follow-up | 1.00 (1.00-1.00) | 1.00 (1.00-1.00) | 1.00 (1.00-1.00) | 1.00 (1.00-1.00) |
| Year of end of record in period (ref final year)<sup>e</sup> | Year 2 | **0.00 (00.00-0.00)** | **0.00 (00.00-0.00)** | 0.28 (0.04-2.14) | **4.67 (1.05-20.71)** |
|  | Year 3 | NA | NA | **1.35 (1.21-1.50)** | **2.47 (2.10-2.90)** |

OR: Odds ratio; 95% CI: 95% confidence interval; ref: Reference category; SMI: Severe Mental Illness; QOF: Quality and Outcomes Framework
a: Multinomial logistic regression comparing patients receiving no screening or receiving complete screening (all six cardiovascular risk factors for each year that the patient is active in the time period) to those who were irregularly screened, with mutual adjustment for all covariates.
b: Exception reported from mental health measures
c: Defined as presence on QOF register for atrial fibrillation, coronary heart disease, hypertension, peripheral artery disease, stroke or diabetes.
d: In period variables measured cross-sectionally, up to the end of the period of interest e: Defined as the year a patient ends follow-up. For the 2014-2018 cohort, year two is 2015-2016, year 3 is 2016-2017 and the final year is 2017-2018. For the 2011-2014 cohort year 2 is 2012-2013 and the final year is 2013-2014. Note: Patients could be present in both time periods. The total number of unique patients is 119,976

In the 2014-2018 period only, older age was associated with lower odds of receiving no screening (OR per 10-year increase in age: 0.86; 95%CI:0.83-0.90), and compared to bipolar disorder, a diagnosis of ‘other psychoses’ was associated with higher odds of receiving no screening (Table 4). In the 2011-2014 period only, compared to White ethnicity, Black ethnicity was associated with higher odds of receiving no screening (OR:1.31; 95%CI:1.03-1.67).

Men were both more likely to have received no screening, and to have always received complete screening than women in both time periods. Men, older patients, those of Asian ethnicity (compared to those of White ethnicity), with a diagnosis of schizophrenia (compared to bipolar disorder), prescribed antipsychotics or mood stabilisers, on a QOF register that incentivised CVD risk factor, and who were never exception reported were more likely to always receive complete screening in all time periods, in unadjusted and adjusted models (Table 3 & 4 and Table S4). Patients of Black (versus White) ethnicity were more likely to always have complete screening in the 2014-2018 period only (OR:1.39; 95%CI:1.19-1.62, Table 4).

Characteristics associated with receipt of screening differed for individual CVD risk factors. For example, in the 2014-2018 period men were more likely to have received regular glucose, cholesterol or smoking screening, but women were more likely to have received regular blood pressure screening. Patients of Black ethnicity were less likely to have received no screening for glucose, cholesterol, and BMI, had similar odds of alcohol and blood pressure screening but were more likely to have received no smoking screening than patients of White ethnicity (Table S5).

### Sensitivity analysis: Investigating the effect of deprivation and follow-up time

Limiting the population to those with available deprivation data or those with complete follow-up for each period did not alter most findings (Table S6-S7). Deprivation was not associated with receiving always complete or no screening, except for in the 2014-2018 period, where those in the most deprived quintile were less likely to have received no screening than those in the least deprived quintile (OR:0.74; 95%CI:0.56-0.99, Table S6).

## Discussion

We found that cardiovascular risk factor screening prevalence varied by individual CVD risk factor, over time, and by patient characteristics. Our findings suggest that patient characteristics and financial incentivisation influence screening prevalence of individual CVD risk factors, the likelihood of receiving screening for all six CVD risk factors annually, and risk of receiving no screening. However, as the patient characteristics associated with increased risk of receiving no screening change over the study periods, it is likely that the groups most at risk of missing screening are dynamic and dependent on which measures are incentivised. Between 2014 and 2018, men, younger patients and those without pre-existing conditions, not prescribed antipsychotics or mood stabilisers, of missing or ‘other’ ethnicity, or with a diagnosis of ‘other psychoses’ had an elevated risk of receiving no screening.

We found a sharp increase in CVD risk factor screening in 2011, most notable for alcohol, BMI, cholesterol and glucose screening, and coinciding with the introduction of incentivisation of all six of the CVD risk factors studied. The subsequent reduction in prevalence of glucose, cholesterol and BMI screening in 2014 coincides with the removal of these measures from incentivisation in England and Northern Ireland, a pattern not seen in the screening prevalence of CVD risk factors which remained incentivised in these countries^22^. This is in line with a recent study by Matias et al, which found a significant reduction in screening for cholesterol and BMI in this period compared to blood pressure in patients with SMI^19^, and studies in the general population which show a decrease in target achievement when incentivisation is withdrawn^25^.

### Strengths and limitations

The large population size allowed us to stratify results by a range of patient characteristics, while the representative nature of CPRD data makes these results generalisable to the UK for this time period, although with notable differences across the four nations of the UK from 2014. Focusing on longitudinal screening at an individual level, as well as screening prevalence at a population level allowed a better understanding of screening practice over time and the identification of groups of patients who are at risk of receiving no screening. Our results highlight the importance of considering regularity and comprehensiveness of screening at an individual level when evaluating screening interventions, rather than reliance on prevalence of screening at a population level.

We required that patients had at least one year of during the study period to reliably capture screening activity. This may mean that patients who are transiently registered, who are less likely to be screened, are not included in our results. Our analysis was exploratory in nature and while we investigated a range of factors that we hypothesised were associated with receipt of CVD risk factor screening in primary care, it is likely that many of these factors interact to produce distinct risk profiles. Furthermore, we did not investigate outcomes of screening practices and therefore cannot determine the impact that screening has on for example stopping smoking, having controlled blood pressure or starting statins, nor on the longer-term health of people with SMI. There is a need for further hypothesis-driven studies to identify groups of patients at risk of missed screening, the impact this has on cardiovascular health and into the effectiveness of physical health checks and CVD risk factor screening in this population.

### Clinical Implications

Given the known cardiovascular health inequalities in those with SMI^3–7^, the finding that only half of patients with SMI had ever received screening for all six CVD risk factors is concerning. Our results suggests that while incentivisation led to an increase in the annual prevalence of screening for individual CVD risk factors, few patients received regular comprehensive screening. In 2023, the proportion of patients with SMI receiving screening for all six CVD risk factors was incentivised for the first time^26^ ^27^. While data collected by NHS England for physical health checks performed in primary or secondary care suggests this has increased the prevalence of screening, ^28^our study highlights the need to consider regularity of screening as well as differences in screening prevalence, the risk of receiving no screening and always receiving complete screening by patient characteristics. We found a low prevalence of screening, and high risk of receiving no screening in those with missing ethnicity, without pre-existing conditions or of younger age. While for those with missing ethnicity, this is likely driven by lack of healthcare contact underpinning both poor ethnicity recording and low levels of screening, for those of younger age or without other physical health conditions it may be driven by additional factors, such as patient and provider perceptions of need. Further interventions are therefore required to improve uptake in these groups.

Incentivisation appears to confer little benefit to those outside of the incentivisation criteria. For example, we found that when cholesterol and glucose screening was incentivised for patients with SMI aged over 40, there was minimal increase in screening in those under 40, so little evidence of halo effects. Likewise, those with ‘other psychoses’ had a lower screening prevalence of all CVD risk factors than patients with schizophrenia or bipolar disorder. The term ‘other psychoses’ covers a range of psychotic diagnoses and symptoms, and is broader than the list included under QOF incentivisation. These findings highlight the careful planning needed when implementing incentivisation so that those falling outside of incentivisation but still at risk are not marginalised, particularly in those with SMI, who may develop multimorbidity at an early age^1^, and where formal diagnosis may be delayed.

People with SMI are at risk of physical health conditions beyond CVD^1^, and of avoidable physical health hospitalisations^29^. Current NHS guidance describes the incentivised CVD risk factors as core to the physical health check in primary care, but recommends a more comprehensive annual review of physical health^30^. We found that only a third of patients ever received screening of all six CVD risk factors within a month of each other suggesting that CVD risk factors may be captured opportunistically. While opportunistic screening results in higher screening prevalence, screening is only a first step. It is important that patients also receive a comprehensive clinical review of physical health so that coordinated actions can be put in place to manage CVD risk, and diagnose and treat conditions beyond CVD. While physical health screening is embedded in primary care, further consideration is warranted as to the provision of physical health checks in mental health services to complement those in primary care and to provide services to those not regularly in contact with their primary care provider. However, this approach is reliant on improved coordination between physical and mental health care providers to avoid duplication for both patient and practitioner, and to ensure transfer of important patient information.

### Conclusions

The low proportion of people receiving regular CVD risk factor screening suggests that people with SMI are not reliably receiving regular comprehensive physical health checks in primary care. Further consideration is warranted as to how incentivisation and other schemes could improve the regularity and comprehensiveness of screening, rather than just the annual screening prevalence, and whether provision of physical health checks in both physical and mental health services may improve up-take. Hypothesis-driven work is required to identify groups of patients most at risk of not receiving CVD risk factor screening so that targeted interventions can be developed, with consideration of these groups in the planning and implementation of future incentivisation and other improvement schemes.

## Data Availability

Data were obtained from a third party and are not publicly available. All data were obtained from Clinical Practice Research Datalink (CPRD). Data is available from CPRD but is subject to protocol approval and ethical review.

## Data sharing statement

Data were obtained from a third party and are not publicly available. All data were obtained from Clinical Practice Research Datalink (CPRD). Data is available from CPRD but is subject to protocol approval and ethical review: https://www.cprd.com/research-applications.

## Acknowledgments

The authors wish to acknowledge the input from the wider collaborators involved in DATAMIND, and particularly the input from lived experience advisors.

## Author contributions

NL wrote the protocol, analysed the data and wrote the first draft of the manuscript. JFH and DPJO had oversight of the data and contributed to the initial study design. CAJ, JFH, AJ, RS, MHI, EB and DPJO commented on the study protocol and helped formulate the original research question. All authors provided input on the interpretation of results and commented on the first draft of the manuscript.

