## Supplementary materials for "Prevalence and patient characteristics associated with cardiovascular disease risk factor screening in UK primary care for people with severe mental illness: An electronic healthcare record study"

### Supplementary material

Table S1: QOF timeline illustrating which physical health risk factors were incentivised during the study period (2004-2018) and after (2019-2024)

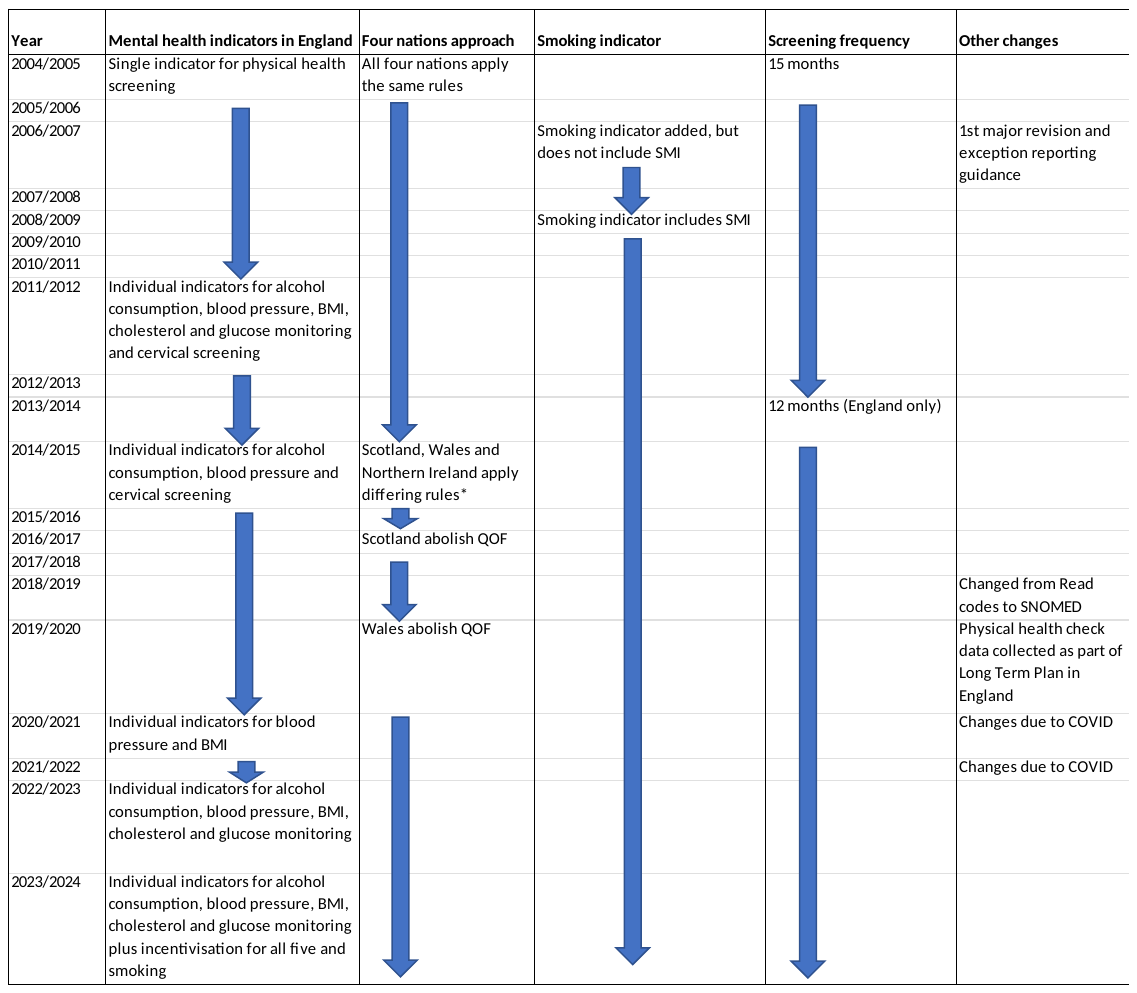

Taken from ^22^ and ^31^. BMI: Body mass index; QOF: Quality and Outcomes Framework

*From 2014: In Wales QOF incentivised glucose, blood pressure and BMI screening in a 15-month period as one indicator and alcohol, smoking and cervical screening in a 15-month period as individual indicators ^32 33^. In Scotland QOF incentivised screening of all six cardiovascular risk factors as individual indicators in a 15-month period ^34^. In Northern Ireland, QOF incentivised blood pressure, smoking and cervical screening in a 15 month period ^35^.

Table S2: Code lists for outcomes, population and covariates

| **Condition** | **Link to code list** |
| --- | --- |
| Severe mental illness | <https://phenotypes.healthdatagateway.org/phenotypes/PH1649/version/3407/detail/> |
| Glucose | <https://phenotypes.healthdatagateway.org/phenotypes/PH1656/version/3422/detail/> |
| Cholesterol | <https://phenotypes.healthdatagateway.org/phenotypes/PH1655/version/3421/detail/> |
| Blood pressure | <https://phenotypes.healthdatagateway.org/phenotypes/PH1654/version/3420/detail/> |
| BMI | <https://phenotypes.healthdatagateway.org/phenotypes/PH1652/version/3404/detail/> |
| Smoking | <https://phenotypes.healthdatagateway.org/phenotypes/PH1651/version/3408/detail/> |
| Alcohol | https://phenotypes.healthdatagateway.org/phenotypes/PH1678/version/3543/detail/ |
| Antipsychotics/mood stabilisers | <https://phenotypes.healthdatagateway.org/phenotypes/PH1650/version/3406/detail/> |
| Exception reporting | https://phenotypes.healthdatagateway.org/phenotypes/PH1679/version/3544/detail/ |

Table S3: Annual prevalence of individual cardiovascular risk factor screening in patients with severe mental illness by financial year, n=216,136

|  | Cholesterol | | Glucose | | Blood pressure | | BMI | | Smoking | | Alcohol | |
| --- | --- | --- | --- | --- | --- | --- | --- | --- | --- | --- | --- | --- |
| Year, (total eligible) | Number screened | Screening prevalence (95%CI) | Number screened | Screening prevalence (95%CI) | Number screened | Screening prevalence (95%CI) | Number screened | Screening prevalence (95%CI) | Number screened | Screening prevalence (95%CI) | Number screened | Screening prevalence (95%CI) |
| 2000-2001 (57,814) | 4,083 | 7.1  (6.9-7.3) | 5,731 | 9.9  (9.7-10.1) | 16,771 | 29.0  (28.64-29.4) | 8,435 | 14.6  (14.3-14.9) | 8,025 | 13.9  (13.6-14.2) | 5,518 | 9.5  (9.3-9.8) |
| 2001-2002 (61,418) | 5,783 | 9.4  (9.2-9.6) | 7,978 | 13.0  (12.7-13.3) | 20,077 | 32.7  (32.3-33.1) | 10,503 | 17.1  (16.8-17.4) | 10,795 | 17.6  (17.3-17.9) | 7,036 | 11.5  (11.2-11.7) |
| 2002-2003 (65,266) | 7,743 | 11.9  (11.6-12.1) | 10,616 | 16.3  (16.0-16.6) | 23,507 | 36.0  (35.7-36.4) | 12,811 | 19.6  (19.3-19.9) | 13,735 | 21.0  (20.7-21.4) | 8,988 | 13.8  (13.5-14.0) |
| 2003-2004 (69,276) | 11,117 | 16.1  (15.8-16.3) | 14,333 | 20.7  (20.4-21.0) | 30,327 | 43.8  (43.4-44.2) | 17,552 | 25.3  (25.0-25.7) | 25,550 | 36.9  (36.5-37.2) | 14,193 | 20.5  (20.2-20.8) |
| 2004-2005 (72,827) | 15,871 | 21.8  (21.5-22.1) | 19,226 | 26.4  (26.1-26.7) | 40,029 | 55.0  (54.6-55.3) | 24,466 | 33.6  (33.3-33.9) | 37,349 | 51.3  (50.9-51.7) | 19,821 | 27.2  (26.9-27.5) |
| 2005-2006 (75,007) | 18,658 | 24.9  (24.6-25.2) | 22,190 | 29.6  (29.3-29.9) | 41,262 | 55.0  (54.7-55.4) | 26,997 | 36.0  (35.7-36.3) | 35,584 | 47.4  (47.1-47.8) | 19,523 | 26.0  (25.7-26.3) |
| 2006-2007 (76,951) | 23,092 | 30.0  (29.7-30.3) | 26,385 | 34.3  (34.0-34.6) | 47,926 | 62.3  (61.9-62.6) | 36,339 | 47.2  (46.9-47.6) | 47,575 | 61.8  (61.5-62.2) | 26,005 | 33.8  (33.5-34.1) |
| 2007-2008 (79,059) | 24,792 | 31.4  (31.0-31.7) | 28,143 | 35.6  (35.3-35.9) | 47,779 | 60.4  (60.1-60.8) | 35,842 | 45.3  (45.0-45.7) | 44,313 | 56.1  (55.7-56.4) | 24,921 | 31.5  (31.2-31.9) |
| 2008-2009 (80,372) | 28,235 | 35.1  (34.8-35.5) | 31,297 | 38.9  (38.6-39.3) | 51,040 | 63.5  (63.2-63.8) | 40,017 | 49.8  (49.4-50.1) | 55,614 | 69.2  (68.9-69.5) | 29,154 | 36.3  (35.9-36.6) |
| 2009-2010 (82,242) | 29,961 | 36.4  (36.1-36.8) | 32,697 | 39.8  (39.4-40.1) | 52,356 | 63.7  (63.3-64.0) | 41,694 | 50.7  (50.4-51.0) | 53,808 | 65.4  (65.1-65.8) | 31,197 | 37.9  (37.6-38.3) |
| 2010-2011 (83,749) | 32,474 | 38.8  (38.5-39.1) | 35,506 | 42.4  (42.1-42.7) | 54,693 | 65.3  (65.0-65.6) | 43,884 | 52.4  (52.1-52.7) | 55,460 | 66.2  (65.9-66.5) | 33,623 | 40.2  (39.8-40.5) |
| 2011-2012 (85,592) | 49,507 | 57.8  (57.5-58.2) | 51,386 | 60.0  (59.7-60.4) | 63,421 | 74.1  (73.8-74.4) | 58,492 | 68.3  (68.0-68.7) | 61,127 | 71.4  (71.1-71.7) | 57,266 | 66.9  (66.6-67.2) |
| 2012-2013 (86,870) | 44,904 | 51.7  (51.4-52.0) | 47,984 | 55.2  (54.9-55.6) | 61,797 | 71.1  (70.8-71.4) | 54,729 | 63.0  (62.7-63.3) | 58,538 | 67.4  (67.1-67.7) | 51,355 | 59.1  (58.8-59.4) |
| 2013-2014 (85,921) | 51,459 | 59.9  (59.6-60.2) | 54,479 | 63.4  (63.1-63.7) | 67,781 | 78.9  (78.6-79.2) | 62,698 | 73.0  (72.7-73.3) | 64,300 | 74.8  (74.5-75.1) | 62,395 | 72.6  (72.3-72.9) |
| 2014-2015 (86,674) | 40,036 | 46.2  (45.9-46.5) | 44,409 | 51.2  (50.9-51.6) | 66,290 | 76.5  (76.2-76.8) | 47,447 | 54.7  (54.4-55.1) | 62,501 | 72.1  (71.8-72.4) | 61,415 | 70.9  (70.6-71.2) |
| 2015-2016 (86482) | 40,002 | 46.3  (45.9-46.6) | 44,632 | 51.6  (51.3-51.9) | 66,034 | 76.4  (76.1-76.6) | 46,546 | 53.8  (53.5-54.2) | 62,107 | 71.8  (71.5-72.1) | 61,194 | 70.8  (70.5-71.1) |
| 2016-2017 (86895) | 39,885 | 45.9  (45.6-46.2) | 44,299 | 51.0  (50.7-51.3) | 66,059 | 76.0  (75.7-76.3) | 46,440 | 53.4  (53.1-53.8) | 61,581 | 70.9  (70.6-71.2) | 60,504 | 69.6  (69.3-69.9) |
| 2017-2018 (80419) | 37,515 | 46.7  (46.3-47.0) | 41,525 | 51.6  (51.3-52.0) | 61,024 | 75.9  (75.6-76.2) | 42,254 | 52.5  (52.2-52.9) | 55,644 | 69.2  (68.9-69.5) | 54,818 | 68.2  (67.8-68.5) |

95%CI: 95% confidence intervals; BMI: body mass index

Table S4: Multinomial logistic regression for the odds ratios of always receiving complete screening or receiving no screening compared to irregular screening in patients with severe mental illness for the period of April 2004 to March 2011, n=106,747

| Reference: Irregular screening | | Complete (n=1,770, 1.66%) | None (n=5,795, 5.43%) |
| --- | --- | --- | --- |
|  | | **OR (95% CI)** | **OR (95% CI)** |
| Age at start of follow-up | Per 10-year increase | **1.23 (1.20-1.27)** | **0.95 (0.91-0.98)** |
| Sex (ref female) | Male | **1.38 (1.24-1.53)** | **1.36 (1.26-1.46)** |
| Ethnicity (ref White) | Asian | **2.36 (1.88-2.97)** | 1.23 (0.97-1.56) |
|  | Black | **1.54 (1.19-1.99)** | **1.50 (1.23-1.84)** |
|  | Mixed | **1.62 (1.07-2.44)** | 1.10 (0.75-1.61) |
|  | Other | **0.55 (0.34-0.88)** | **2.71 (2.15-3.42)** |
|  | Missing | **0.57 (0.48-0.67)** | **4.96 (4.16-5.90)** |
| Country (ref England) | Northern Ireland | 0.70 (0.32-1.51) | **0.41 (0.30-0.57)** |
|  | Scotland | 1.11 (0.82-1.50) | 0.55 (0.46-0.66) |
|  | Wales | 1.03 (0.75-1.40) | 0.47 (0.37-0.59) |
| SMI diagnosis (ref bipolar) | Schizophrenia | **1.22 (1.08-1.37)** | **1.25 (1.14-1.37)** |
|  | Other psychoses | 0.89 (0.78-1.01) | **1.16 (1.05-1.29)** |
| In period variables | Exception reported | **0.55 (0.47-0.65)** | 0.98 (0.80-1.19) |
|  | Other QOF register^a^ | **8.37 (6.74-10.41)** | **0.31 (0.28-0.33)** |
|  | On antipsychotics/mood stabilisers | **1.29 (1.14-1.46)** | **0.16 (0.14-0.19)** |
|  | Years since diagnosis | 1.00 (1.00-1.00) | 1.00 (1.00-1.00) |
|  | Years since registration | 1.00 (1.00-1.00) | 1.00 (1.00-1.00) |
|  | Years of follow-up | 1.00 (1.00-1.00) | 1.00 (1.00-1.00) |
| Year of end of record in period (ref 2010-2011) | 2005-2006 | **5.15 (1.30-20.41)** | **4.78 (1.13-20.22)** |
|  | 2006-2007 | 1.05 (0.85-1.30) | **4.91 (4.13-5.84)** |
|  | 2007-2008 | 0.90 (0.73-1.11) | **2.24 (1.89-2.66)** |
|  | 2008-2009 | 0.86 (0.69-1.07) | **2.43 (1.29-4.58)** |
|  | 2009-2010 | **0.76 (0.61-0.94)** | **1.26 (1.04-1.53)** |

OR: Odds ratio; 95% CI: 95% confidence interval; ref: Reference category; SMI: Severe Mental Illness; QOF: Quality and Outcomes Framework

a: Defined as presence on QOF register for atrial fibrillation, coronary heart disease, hypertension, peripheral artery disease, stroke or diabetes.

Table S5: Multinomial logistic regression for the odds ratios of always receiving complete screening or receiving no screening compared to irregular screening for individual cardiovascular risk factors in people with severe mental illness, April 2011-March 2018, n=119,976

| Reference: Irregular screening | |  | April 2011 – March 2014 (n=85,274) | | April 2014 – March 2018 (n=94,216) | |
| --- | --- | --- | --- | --- | --- | --- |
|  | |  | Complete (n=12,616, 14.79%) | None (n=3,204, 3.76%) | Complete (n=12,616, 14.79%) | None (n=3,204, 3.76%) |
|  | |  | OR (95% CI) | OR (95% CI) | OR (95% CI) | OR (95% CI) |
| Blood pressure | **Age at start of follow-up** | Per 10-year increase | 1.29 (1.27-1.31) | 0.92 (0.90-0.96) | 1.27 (1.26-1.29) | 0.84 (0.83-0.87) |
|  | **Sex (ref female)** | Male | 0.90 (0.87-0.93) | 1.47 (1.39-1.56) | 0.87 (0.84-0.89) | 1.51 (1.41-1.60) |
|  | **Ethnicity (ref white)** | Asian | 1.34 (1.23-1.47) | 0.86 (0.74-1.00) | 1.40 (1.27-1.54) | 0.85 (0.72-1.00) |
|  |  | Black | 1.27 (1.17-1.38) | 0.97 (0.83-1.15) | 1.20 (1.10-1.30) | 1.06 (0.93-1.21) |
|  |  | Mixed | 1.04 (0.90-1.21) | 1.00 (0.80-1.24) | 1.04 (0.92-1.18) | 1.01 (0.80-1.27) |
|  |  | Other | 0.82 (0.73-0.92) | 1.55 (1.31-1.84) | 0.95 (0.85-1.07) | 1.27 (1.06-1.53) |
|  |  | Missing | 0.74 (0.69-0.78) | 2.62 (2.32-2.96) | 0.79 (0.75-0.84) | 1.53 (1.42-1.66) |
|  | **Country (ref England)** | NI | 0.89 (0.74-1.07) | 0.45 (0.37-0.55) | 0.44 (0.34-0.56) | 0.69 (0.52-0.90) |
|  |  | Scotland | 1.05 (0.96-1.16) | 0.90 (0.79-1.03) | 0.39 (0.34-0.44) | 1.87 (1.65-2.12) |
|  |  | Wales | 0.88 (0.80-0.97) | 0.64 (0.54-0.76) | 0.43 (0.39-0.47) | 1.21 (1.04-1.41) |
|  | **SMI diagnosis (ref bipolar disorder)** | Schizophrenia | 1.04 (1.00-1.09) | 1.03 (0.96-1.11) | 0.99 (0.95-1.04) | 0.99 (0.91-1.07) |
|  |  | Other psychoses | 0.89 (0.86-0.93) | 1.17 (1.09-1.26) | 0.89 (0.85-0.92) | 1.18 (1.10-1.27) |
|  | **In period variables** | **Exception reported** | 0.42 (0.40-0.43) | 1.69 (1.54-1.86) | 0.37 (0.36-0.39) | 1.60 (1.51-1.71) |
|  |  | **Other QOF register^a^** | 2.13 (2.05-2.21) | 0.58 (0.54-0.62) | 2.01 (1.94-2.08) | 0.59 (0.55-0.62) |
|  |  | **On antipsychotics/mood stabilisers** | 1.73 (1.65-1.81) | 0.32 (0.30-0.35) | 2.07 (1.98-2.16) | 0.40 (0.38-0.43) |
|  |  | **Time since diagnosis** | 1.00 (1.00-1.00) | 1.00 (1.00-1.00) | 1.00 (1.00-1.00) | 1.00 (1.00-1.00) |
|  |  | **Time since registration** | 1.00 (1.00-1.00) | 1.00 (1.00-1.00) | 1.00 (1.00-1.00) | 1.00 (1.00-1.00) |
|  |  | **Years of follow-up** | 1.00 (1.00-1.00) | 1.00 (1.00-1.00) | 1.00 (1.00-1.00) | 1.00 (1.00-1.00) |
|  | **Year of end of follow-up in period (ref final year)** | Year 2 | 0.66 (0.17-2.52) | 0.00 (0.00-0.00) | 1.01 (0.46-2.19) | 2.78 (0.93-8.33) |
|  |  | Year 3 | NA | NA | 1.52 (1.38-1.66) | 2.27 (2.06-2.51) |
| Glucose | **Age at start of follow-up** | **Per 10-year increase** | 1.28 (1.27-1.29) | 0.73 (0.71-0.75) | 1.21 (1.20-1.22) | 0.80 (0.79-0.82) |
|  | **Sex (ref female)** | Male | 1.08 (1.05-1.12) | 1.19 (1.14-1.23) | 1.12 (1.08-1.16) | 1.25 (1.21-1.30) |
|  | **Ethnicity (ref white)** | Asian | 1.41 (1.29-1.54) | 0.80 (0.72-0.89) | 1.85 (1.71-2.01) | 0.67 (0.60-0.74) |
|  |  | Black | 1.18 (1.08-1.28) | 0.97 (0.88-1.07) | 1.45 (1.33-1.57) | 0.89 (0.82-0.97) |
|  |  | Mixed | 1.02 (0.88-1.18) | 1.16 (1.00-1.36) | 1.16 (1.00-1.35) | 1.04 (0.91-1.19) |
|  |  | Other | 0.89 (0.78-1.01) | 1.48 (1.29-1.70) | 1.04 (0.90-1.20) | 1.13 (0.99-1.28) |
|  |  | Missing | 0.83 (0.78-0.88) | 1.84 (1.69-2.01) | 0.84 (0.79-0.89) | 1.41 (1.33-1.49) |
|  | **Country (ref England)** | NI | 1.20 (0.91-1.60) | 0.53 (0.43-0.66) | 0.98 (0.80-1.21) | 0.80 (0.66-0.98) |
|  |  | Scotland | 1.20 (1.07-1.34) | 0.94 (0.83-1.07) | 1.20 (1.06-1.36) | 1.05 (0.95-1.16) |
|  |  | Wales | 0.94 (0.84-1.05) | 0.75 (0.67-0.85) | 1.24 (1.13-1.36) | 0.65 (0.59-0.71) |
|  | **SMI diagnosis (ref bipolar disorder)** | Schizophrenia | 1.06 (1.02-1.11) | 1.02 (0.97-1.08) | 1.17 (1.11-1.22) | 1.07 (1.02-1.12) |
|  |  | Other psychoses | 0.88 (0.84-0.91) | 1.23 (1.17-1.30) | 0.95 (0.91-1.00) | 1.26 (1.21-1.32) |
|  | **In period variables** | **Exception reported** | 0.46 (0.44-0.48) | 1.76 (1.66-1.87) | 0.61 (0.58-0.64) | 1.78 (1.70-1.86) |
|  |  | **Other QOF register^a^** | 1.77 (1.70-1.85) | 0.63 (0.60-0.66) | 2.32 (2.21-2.44) | 0.66 (0.64-0.69) |
|  |  | **On antipsychotics/mood stabilisers** | 1.90 (1.80-1.99) | 0.38 (0.36-0.40) | 1.63 (1.55-1.72) | 0.45 (0.43-0.47) |
|  |  | **Time since diagnosis** | 1.00 (1.00-1.00) | 1.00 (1.00-1.00) | 1.00 (1.00-1.00) | 1.00 (1.00-1.00) |
|  |  | **Time since registration** | 1.00 (1.00-1.00) | 1.00 (1.00-1.00) | 1.00 (1.00-1.00) | 1.00 (1.00-1.00) |
|  |  | **Years of follow-up** | 1.00 (1.00-1.00) | 1.00 (1.00-1.00) | 1.00 (1.00-1.00) | 1.00 (1.00-1.00) |
|  | **Year of end of follow-up in period (ref final year)** | Year 2 | 0.41 (0.09-1.93) | 0.00 (00.00-0.00) | 0.90 (0.29-2.80) | 3.08 (1.29-7.39) |
|  |  | Year 3 | NA | NA | 1.56 (1.45-1.69) | 1.64 (1.52-1.78) |
| Cholesterol | **Age at start of follow-up** | Per 10-year increase | 1.23 (1.22-1.24) | 0.68 (0.66-0.69) | 1.16 (1.15-1.18) | 0.78 (0.77-0.80) |
|  | **Sex (ref female)** | Male | 1.16 (1.12-1.20) | 0.99 (0.95-1.03) | 1.21 (1.16-1.25) | 0.97 (0.94-1.01) |
|  | **Ethnicity (ref white)** | Asian | 1.34 (1.22-1.48) | 0.80 (0.72-0.89) | 1.72 (1.58-1.88) | 0.63 (0.57-0.70) |
|  |  | Black | 1.08 (0.98-1.18) | 0.90 (0.82-1.00) | 1.42 (1.30-1.54) | 0.82 (0.76-0.89) |
|  |  | Mixed | 0.98 (0.85-1.14) | 1.10 (0.94-1.28) | 1.22 (1.04-1.43) | 1.05 (0.92-1.19) |
|  |  | Other | 0.87 (0.77-0.99) | 1.49 (1.31-1.69) | 0.94 (0.80-1.09) | 1.18 (1.05-1.33) |
|  |  | Missing | 0.78 (0.73-0.83) | 1.80 (1.66-1.96) | 0.85 (0.79-0.92) | 1.51 (1.42-1.59) |
|  | **Country (ref England)** | Northern Ireland | 1.14 (0.85-1.52) | 0.57 (0.47-0.69) | 0.99 (0.73-1.33) | 0.84 (0.69-1.03) |
|  |  | Scotland | 1.32 (1.18-1.47) | 0.92 (0.83-1.03) | 1.46 (1.24-1.71) | 1.08 (0.96-1.21) |
|  |  | Wales | 0.87 (0.78-0.98) | 0.77 (0.69-0.87) | 0.80 (0.69-0.92) | 0.91 (0.81-1.02) |
|  | **SMI diagnosis (ref bipolar disorder)** | Schizophrenia | 1.02 (0.98-1.06) | 0.95 (0.90-0.99) | 1.12 (1.07-1.17) | 0.92 (0.88-0.96) |
|  |  | Other psychoses | 0.87 (0.84-0.91) | 1.25 (1.19-1.31) | 0.95 (0.90-0.99) | 1.23 (1.18-1.28) |
|  | **In period variables** | **Exception reported** | 0.47 (0.45-0.49) | 1.78 (1.69-1.88) | 0.61 (0.58-0.64) | 1.70 (1.63-1.77) |
|  |  | **Other QOF register^a^** | 1.54 (1.48-1.60) | 0.65 (0.62-0.68) | 2.15 (2.03-2.27) | 0.69 (0.67-0.72) |
|  |  | **On antipsychotics/mood stabilisers** | 1.64 (1.57-1.73) | 0.37 (0.35-0.39) | 1.46 (1.38-1.55) | 0.45 (0.43-0.47) |
|  |  | **Time since diagnosis** | 1.00 (1.00-1.00) | 1.00 (1.00-1.00) | 1.00 (1.00-1.00) | 1.00 (1.00-1.00) |
|  |  | **Time since registration** | 1.00 (1.00-1.00) | 1.00 (1.00-1.00) | 1.00 (1.00-1.00) | 1.00 (1.00-1.00) |
|  |  | **Years of follow-up** | 1.00 (1.00-1.00) | 1.00 (1.00-1.00) | 1.00 (1.00-1.00) | 1.00 (1.00-1.00) |
|  | **Year of end of follow-up in period (ref final year)** | Year 2 | 0.34 (0.08-1.49) | 0.26 (0.02-2.90) | 0.52 (0.13-2.01) | 3.22 (1.34-7.78) |
|  |  | Year 3 | NA | NA | 1.50 (1.38-1.63) | 1.73 (1.60-1.88) |
| BMI | **Age at start of follow-up** | Per 10-year increase | 1.06 (1.05-1.07) | 1.01 (0.99-1.03) | 1.07 (1.05-1.08) | 0.98 (0.97-1.00) |
|  | **Sex (ref female)** | Male | 0.92 (0.89-0.95) | 1.16 (1.11-1.22) | 1.02 (0.98-1.06) | 1.18 (1.14-1.23) |
|  | **Ethnicity (ref white)** | Asian | 1.25 (1.13-1.38) | 0.88 (0.77-1.00) | 1.39 (1.24-1.55) | 0.77 (0.68-0.87) |
|  |  | Black | 1.10 (1.02-1.20) | 0.91 (0.78-1.06) | 1.24 (1.11-1.39) | 0.85 (0.77-0.94) |
|  |  | Mixed | 1.00 (0.87-1.15) | 0.78 (0.63-0.97) | 0.95 (0.83-1.10) | 0.90 (0.78-1.03) |
|  |  | Other | 0.78 (0.68-0.89) | 1.53 (1.32-1.78) | 0.82 (0.71-0.95) | 1.38 (1.20-1.58) |
|  |  | Missing | 0.75 (0.71-0.80) | 2.36 (2.14-2.60) | 0.82 (0.77-0.88) | 1.79 (1.68-1.90) |
|  | **Country (ref England)** | Northern Ireland | 0.91 (0.69-1.21) | 0.49 (0.40-0.61) | 1.08 (0.82-1.42) | 0.68 (0.51-0.89) |
|  |  | Scotland | 1.10 (0.98-1.24) | 0.99 (0.87-1.13) | 1.09 (0.95-1.25) | 0.99 (0.88-1.11) |
|  |  | Wales | 0.83 (0.74-0.93) | 0.69 (0.60-0.79) | 1.05 (0.94-1.18) | 0.58 (0.51-0.67) |
|  | **SMI diagnosis (ref bipolar disorder)** | Schizophrenia | 1.12 (1.08-1.17) | 0.97 (0.91-1.03) | 1.22 (1.16-1.27) | 0.95 (0.91-1.00) |
|  |  | Other psychoses | 0.89 (0.86-0.93) | 1.13 (1.06-1.21) | 0.99 (0.95-1.04) | 1.24 (1.19-1.30) |
|  | **In period variables** | **Exception reported** | 0.40 (0.38-0.42) | 1.75 (1.64-1.88) | 0.56 (0.54-0.59) | 1.60 (1.53-1.68) |
|  |  | **Other QOF register^a^** | 1.64 (1.58-1.70) | 0.59 (0.55-0.62) | 1.99 (1.90-2.08) | 0.68 (0.65-0.70) |
|  |  | **On antipsychotics/mood stabilisers** | 1.81 (1.73-1.89) | 0.34 (0.32-0.36) | 1.59 (1.51-1.67) | 0.50 (0.48-0.53) |
|  |  | **Time since diagnosis** | 1.00 (1.00-1.00) | 1.00 (1.00-1.00) | 1.00 (1.00-1.00) | 1.00 (1.00-1.00) |
|  |  | **Time since registration** | 1.00 (1.00-1.00) | 1.00 (1.00-1.00) | 1.00 (1.00-1.00) | 1.00 (1.00-1.00) |
|  |  | **Years of follow-up** | 1.00 (1.00-1.00) | 1.00 (1.00-1.00) | 1.00 (1.00-1.00) | 1.00 (1.00-1.00) |
|  | **Year of end of follow-up in period (ref final year)** | Year 2 | 4.81 (0.47-48.80) | 5.62 (0.46-68.08) | 1.08 (0.38-3.09) | 2.66 (1.10-6.46) |
|  |  | Year 3 | NA | NA | 1.47 (1.36-1.60) | 1.83 (1.69-1.99) |
| Alcohol | **Age at start of follow-up** | **Per 10-year increase** | 1.07 (1.06-1.08) | 1.00 (0.98-1.02) | 1.12 (1.09-1.13) | 0.97 (0.95-0.99) |
|  | **Sex (ref female)** | Male | 1.07 (1.03-1.10) | 1.04 (1.00-1.10) | 1.01 (0.98-1.04) | 1.05 (1.00-1.10) |
|  | **Ethnicity (ref white)** | Asian | 1.23 (1.11-1.35) | 0.93 (0.81-1.07) | 1.33 (1.19-1.48) | 0.74 (0.63-0.86) |
|  |  | Black | 1.04 (0.95-1.13) | 0.84 (0.72-0.97) | 1.11 (1.00-1.22) | 0.92 (0.81-1.03) |
|  |  | Mixed | 0.93 (0.80-1.07) | 0.91 (0.72-1.13) | 1.03 (0.90-1.17) | 0.94 (0.76-1.15) |
|  |  | Other | 0.81 (0.71-0.92) | 1.57 (1.34-1.84) | 0.95 (0.82-1.10) | 1.35 (1.13-1.61) |
|  |  | Missing | 0.74 (0.70-0.79) | 2.25 (2.04-2.48) | 0.80 (0.75-0.85) | 1.58 (1.47-1.70) |
|  | **Country (ref England)** | Northern Ireland | 0.79 (0.57-1.09) | 0.53 (0.41-0.68) | 0.27 (0.20-0.35) | 0.48 (0.38-0.61) |
|  |  | Scotland | 1.13 (0.99-1.29) | 0.94 (0.82-1.08) | 0.21 (0.18-0.25) | 1.72 (1.48-2.00) |
|  |  | Wales | 0.75 (0.65-0.86) | 0.67 (0.59-0.77) | 0.19 (0.16-0.23) | 0.97 (0.85-1.11) |
|  | **SMI diagnosis (ref bipolar disorder)** | Schizophrenia | 1.14 (1.09-1.19) | 0.79 (0.74-0.84) | 1.13 (1.09-1.19) | 0.79 (0.73-0.85) |
|  |  | Other psychoses | 0.95 (0.91-0.99) | 1.09 (1.02-1.16) | 0.95 (0.91-0.99) | 1.15 (1.08-1.23) |
|  | **In period variables** | **Exception reported** | 0.40 (0.38-0.42) | 1.45 (1.35-1.55) | 0.33 (0.31-0.35) | 1.08 (1.01-1.15) |
|  |  | **Other QOF register^a^** | 1.46 (1.41-1.52) | 0.60 (0.56-0.63) | 1.44 (1.39-1.49) | 0.65 (0.62-0.69) |
|  |  | **On antipsychotics/mood stabilisers** | 1.72 (1.65-1.80) | 0.29 (0.28-0.31) | 2.27 (2.16-2.38) | 0.37 (0.35-0.39) |
|  |  | **Time since diagnosis** | 1.00 (1.00-1.00) | 1.00 (1.00-1.00) | 1.00 (1.00-1.00) | 1.00 (1.00-1.00) |
|  |  | **Time since registration** | 1.00 (1.00-1.00) | 1.00 (1.00-1.00) | 1.00 (1.00-1.00) | 1.00 (1.00-1.00) |
|  |  | **Years of follow-up** | 1.00 (1.00-1.00) | 1.00 (1.00-1.00) | 1.00 (1.00-1.00) | 1.00 (1.00-1.00) |
|  | **Year of end of follow-up in period (ref final year)** | Year 2 | 0.79 (0.26-2.42) | 1.82 (0.28-11.71) | 0.59 (0.25-1.37) | 1.54 (0.55-4.32) |
|  |  | Year 3 | NA | NA | 1.43 (1.31-1.56) | 2.47 (2.24-2.73) |
| Smoking | **Age at start of follow-up** | Per 10-year increase | 0.96 (0.95-0.97) | 1.16 (1.14-1.18) | 0.91 (0.90-0.92) | 1.15 (1.13-1.17) |
|  | **Sex (ref female)** | Male | 1.09 (1.05-1.12) | 0.92 (0.88-0.97) | 1.25 (1.21-1.29) | 0.98 (0.93-1.03) |
|  | **Ethnicity (ref white)** | Asian | 0.75 (0.69-0.83) | 1.32 (1.15-1.50) | 0.66 (0.60-0.72) | 1.04 (0.91-1.19) |
|  |  | Black | 0.81 (0.75-0.87) | 1.10 (0.95-1.27) | 0.79 (0.72-0.87) | 1.18 (1.05-1.33) |
|  |  | Mixed | 0.87 (0.77-0.99) | 0.84 (0.64-1.09) | 0.95 (0.85-1.06) | 0.97 (0.78-1.21) |
|  |  | Other | 0.77 (0.68-0.89) | 1.67 (1.39-2.00) | 0.87 (0.78-0.98) | 1.31 (1.09-1.58) |
|  |  | Missing | 0.76 (0.71-0.80) | 2.18 (1.95-2.45) | 0.82 (0.78-0.87) | 1.55 (1.44-1.67) |
|  | **Country (ref England)** | Northern Ireland | 0.98 (0.75-1.26) | 0.46 (0.36-0.58) | 0.32 (0.24-0.44) | 0.62 (0.49-0.79) |
|  |  | Scotland | 1.19 (1.07-1.33) | 0.67 (0.58-0.78) | 0.35 (0.30-0.40) | 1.11 (0.98-1.25) |
|  |  | Wales | 0.96 (0.86-1.08) | 0.58 (0.49-0.68) | 0.36 (0.32-0.41) | 0.72 (0.63-0.84) |
|  | **SMI diagnosis (ref bipolar disorder)** | Schizophrenia | 1.20 (1.16-1.25) | 0.94 (0.88-1.00) | 1.27 (1.22-1.32) | 0.88 (0.83-0.95) |
|  |  | Other psychoses | 0.95 (0.92-0.99) | 1.00 (0.93-1.07) | 0.99 (0.95-1.02) | 1.11 (1.05-1.18) |
|  | **In period variables** | **Exception reported** | 0.64 (0.62-0.67) | 1.28 (1.19-1.38) | 0.75 (0.72-0.78) | 1.21 (1.14-1.28) |
|  |  | **Other QOF register^a^** | 1.68 (1.62-1.74) | 0.61 (0.57-0.65) | 1.52 (1.46-1.57) | 0.64 (0.61-0.68) |
|  |  | **On antipsychotics/mood stabilisers** | 1.52 (1.46-1.59) | 0.44 (0.41-0.48) | 1.58 (1.52-1.65) | 0.60 (0.56-0.63) |
|  |  | **Time since diagnosis** | 1.00 (1.00-1.00) | 1.00 (1.00-1.00) | 1.00 (1.00-1.00) | 1.00 (1.00-1.00) |
|  |  | **Time since registration** | 1.00 (1.00-1.00) | 1.00 (1.00-1.00) | 1.00 (1.00-1.00) | 1.00 (1.00-1.00) |
|  |  | **Years of follow-up** | 1.00 (1.00-1.00) | 1.00 (1.00-1.00) | 1.00 (1.00-1.00) | 1.00 (1.00-1.00) |
|  | **Year of end of follow-up in period (ref final year)** | Year 2 | 0.38 (0.12-1.22) | 3.90 (0.83-18.25) | 0.64 (0.28-1.45) | 2.10 (0.73-6.07) |
|  |  | Year 3 | NA | NA | 1.56 (1.46-1.67) | 2.43 (2.19-2.69) |

OR: Odds ratio; 95% CI: 95% confidence interval; ref: Reference category; SMI: Severe Mental Illness; QOF: Quality and Outcomes Framework; BMI: body mass index

a: Defined as presence on QOF register for atrial fibrillation, coronary heart disease, hypertension, peripheral artery disease, stroke or diabetes.

Table S6: Multinomial logistic regression for the odds ratios of always receiving complete screening or receiving no screening compared to irregular screening of cardiovascular risk factors in patients with severe mental illness with available area-based deprivation (IMD) data (England only), n=40,264

|  |  | 2011-2014 (n=29,548) | | 2014-2018 (n=28,762) | |
| --- | --- | --- | --- | --- | --- |
| Reference: Irregular screening | | **Complete**  **(n=4,346, 14.71%)** | **None**  **(n=1,078, 3.65%)** | **Complete (n=2,437, 8.47%)** | **None**  **(n=583, 2.03%)** |
|  |  | **OR (95% CI)** | **OR (95% CI)** | **OR (95% CI)** | **OR (95% CI)** |
| Age at start of follow-up | Per 10-year increase | 1.21 (1.17-1.23) | 0.97 (0.90-1.03) | 1.16 (1.13-1.20) | 0.90 (0.84-0.98) |
| Sex (ref female) | Male | 1.22 (1.14-1.31) | 1.28 (1.11-1.47) | 1.32 (1.21-1.44) | 1.48 (1.22-1.81) |
| Ethnicity (ref White) | Asian | 1.26 (1.05-1.51) | 1.05 (0.72-1.54) | 1.56 (1.23-1.98) | 0.73 (0.47-1.15) |
|  | Black | 0.84 (0.68-1.03) | 1.16 (0.81-1.67) | 1.20 (0.95-1.53) | 0.99 (0.67-1.47) |
|  | Mixed | 0.86 (0.63-1.18) | 0.83 (0.42-1.62) | 0.91 (0.61-1.37) | 0.71 (0.33-1.53) |
|  | Other | 0.83 (0.67-1.04) | 2.46 (1.69-3.60) | 0.93 (0.66-1.30) | 1.13 (0.66-1.96) |
|  | Missing | 0.67 (0.57-0.80) | 4.45 (3.21-6.16) | 0.73 (0.61-0.86) | 2.24 (1.77-2.84) |
| SMI diagnosis (ref bipolar disorder) | Schizophrenia | 1.26 (1.15-1.37) | 0.91 (0.75-1.09) | 1.24 (1.10-1.39) | 0.88 (0.69-1.13) |
|  | Other psychoses | 0.91 (0.82-1.00) | 1.08 (0.90-1.29) | 0.97 (0.86-1.09) | 1.19 (0.95-1.50) |
| In period variables | **Exception reported** | 0.36 (0.31-0.41) | 1.24 (0.93-1.64) | 0.47 (0.41-0.53) | 1.56 (1.26-1.93) |
|  | **Other QOF^a^** | 1.95 (1.77-2.16) | 0.34 (0.29-0.40) | 2.96 (2.54-3.45) | 0.32 (0.26-0.39) |
|  | **On antipsychotics/mood stabilisers** | 1.84 (1.64-2.06) | 0.16 (0.14-0.20) | 2.09 (1.82-2.41) | 0.22 (0.17-0.27) |
|  | **Time since diagnosis** | 1.00 (1.00-1.00) | 1.00 (1.00-1.00) | 1.00 (1.00-1.00) | 1.00 (1.00-1.00) |
|  | **Time since registration** | 1.00 (1.00-1.00) | 1.00 (1.00-1.00) | 1.00 (1.00-1.00) | 1.00 (1.00-1.00) |
|  | **Years of follow-up** | 1.00 (1.00-1.00) | 1.00 (1.00-1.00) | 1.00 (1.00-1.00) | 1.00 (1.00-1.00) |
| Year of end of follow-up in period (ref final year) | Year 2 | 0.00 (00.00-0.00) | 0.00 (00.00-0.00) | 0.00 (00.00-0.00) | 8.06 (0.43-150.80) |
|  | Year 3 | NA | NA | 1.40 (1.16-1.68) | 2.81 (2.12-3.73) |
| Index of multiple deprivation quintile (ref 1 - least deprived) | 2 | 1.01 (0.85-1.21) | 1.00 (0.77-1.31) | 1.08 (0.86-1.35) | 0.94 (0.70-1.26) |
|  | 3 | 1.05 (0.88-1.26) | 1.03 (0.75-1.40) | 1.21 (0.94-1.54) | 0.80 (0.60-1.06) |
|  | 4 | 1.25 (1.05-1.50) | 0.96 (0.61-1.52) | 1.37 (1.07-1.77) | 0.75 (0.56-1.01) |
|  | 5 – Most deprived | 1.27 (1.05-1.54) | 1.16 (0.81-1.65) | 1.61 (1.23-2.09) | 0.74 (0.56-0.99) |

OR: Odds ratio; 95% CI: 95% confidence interval; ref: Reference category; SMI: Severe Mental Illness; QOF: Quality and Outcomes Framework. a: Defined as presence on QOF register for atrial fibrillation, coronary heart disease, hypertension, peripheral artery disease, stroke or diabetes.

Table S7: Multinomial logistic regression for the odds ratios for always receiving complete screening or receiving no screening compared to irregular screening for cardiovascular risk factors in patients active for the full duration of each time period, n=83,838

|  |  | 2011-2014 (66,816) | | 2014-2018 (61,003) | |
| --- | --- | --- | --- | --- | --- |
| Reference: Irregular screening | | **Complete**  **(n=9743, 14.58%)** | **None**  **(n=1742, 2.61%))** | **Complete**  **(n=4708, 7.72%)** | **None**  **(n=1120, 1.84%)** |
|  |  | **OR (95% CI)** | **OR (95% CI)** | **OR (95% CI)** | **OR (95% CI)** |
| Age at start of follow-up | Per 10-year increase | 1.20 (1.17-1.21) | 0.94 (0.90-0.99) | 1.20 (1.17-1.23) | 0.87 (0.82-0.92) |
| Sex (ref female) | Male | 1.24 (1.18-1.30) | 1.30 (1.17-1.45) | 1.37 (1.28-1.46) | 1.47 (1.28-1.68) |
| Ethnicity (ref white) | Asian | 1.27 (1.09-1.46) | 0.92 (0.66-1.28) | 1.85 (1.54-2.23) | 0.89 (0.59-1.35) |
|  | Black | 0.89 (0.77-1.03) | 1.30 (0.96-1.77) | 1.34 (1.10-1.63) | 1.46 (1.08-1.96) |
|  | Mixed | 0.76 (0.61-0.95) | 1.10 (0.68-1.77) | 1.16 (0.87-1.54) | 0.75 (0.40-1.42) |
|  | Other | 0.78 (0.64-0.96) | 2.43 (1.80-3.27) | 0.89 (0.65-1.20) | 1.76 (1.17-2.63) |
|  | Missing | 0.68 (0.62-0.76) | 3.69 (3.19-4.28) | 0.71 (0.62-0.82) | 2.51 (2.12-2.97) |
| Country (ref England) | NI | 1.16 (0.76-1.76) | 0.48 (0.33-0.70) | 0.66 (0.40-1.08) | 0.79 (0.47-1.31) |
|  | Scotland | 1.61 (1.34-1.94) | 0.96 (0.77-1.21) | 1.23 (0.95-1.59) | 2.39 (1.92-2.97) |
|  | Wales | 0.85 (0.70-1.04) | 0.79 (0.59-1.05) | 0.54 (0.42-0.69) | 1.38 (0.98-1.94) |
| SMI diagnosis (ref bipolar disorder) | Schizophrenia | 1.29 (1.21-1.36) | 0.98 (0.85-1.13) | 1.38 (1.28-1.50) | 1.04 (0.86-1.25) |
|  | Other psychoses | 0.95 (0.90-1.01) | 1.21 (1.06-1.38) | 1.02 (0.94-1.11) | 1.36 (1.15-1.61) |
| In period variables | **Exception reported** | 0.39 (0.36-0.42) | 1.61 (1.41-1.85) | 0.50 (0.46-0.54) | 1.61 (1.38-1.87) |
|  | **Other QOF register^a^** | 2.03 (1.90-2.18) | 0.30 (0.27-0.34) | 3.97 (3.45-4.57) | 0.27 (0.23-0.31) |
|  | **On antipsychotics/mood stabilisers** | 1.98 (1.83-2.13) | 0.18 (0.16-0.20) | 2.01 (1.81-2.24) | 0.19 (0.16-0.22) |
|  | **Time since diagnosis** | 1.00 (1.00-1.00) | 1.00 (1.00-1.00) | 1.00 (1.00-1.00) | 1.00 (1.00-1.00) |
|  | **Time since registration** | 1.00 (1.00-1.00) | 1.00 (1.00-1.00) | 1.00 (1.00-1.00) | 1.00 (1.00-1.00) |

OR: Odds ratio; 95% CI: 95% confidence interval; ref: Reference category; SMI: Severe Mental Illness; QOF: Quality and Outcomes Framework.

a: Defined as presence on QOF register for atrial fibrillation, coronary heart disease, hypertension, peripheral artery disease, stroke or diabetes.

Figure S1: Study population flow diagram, detailing inclusion and exclusion criteria

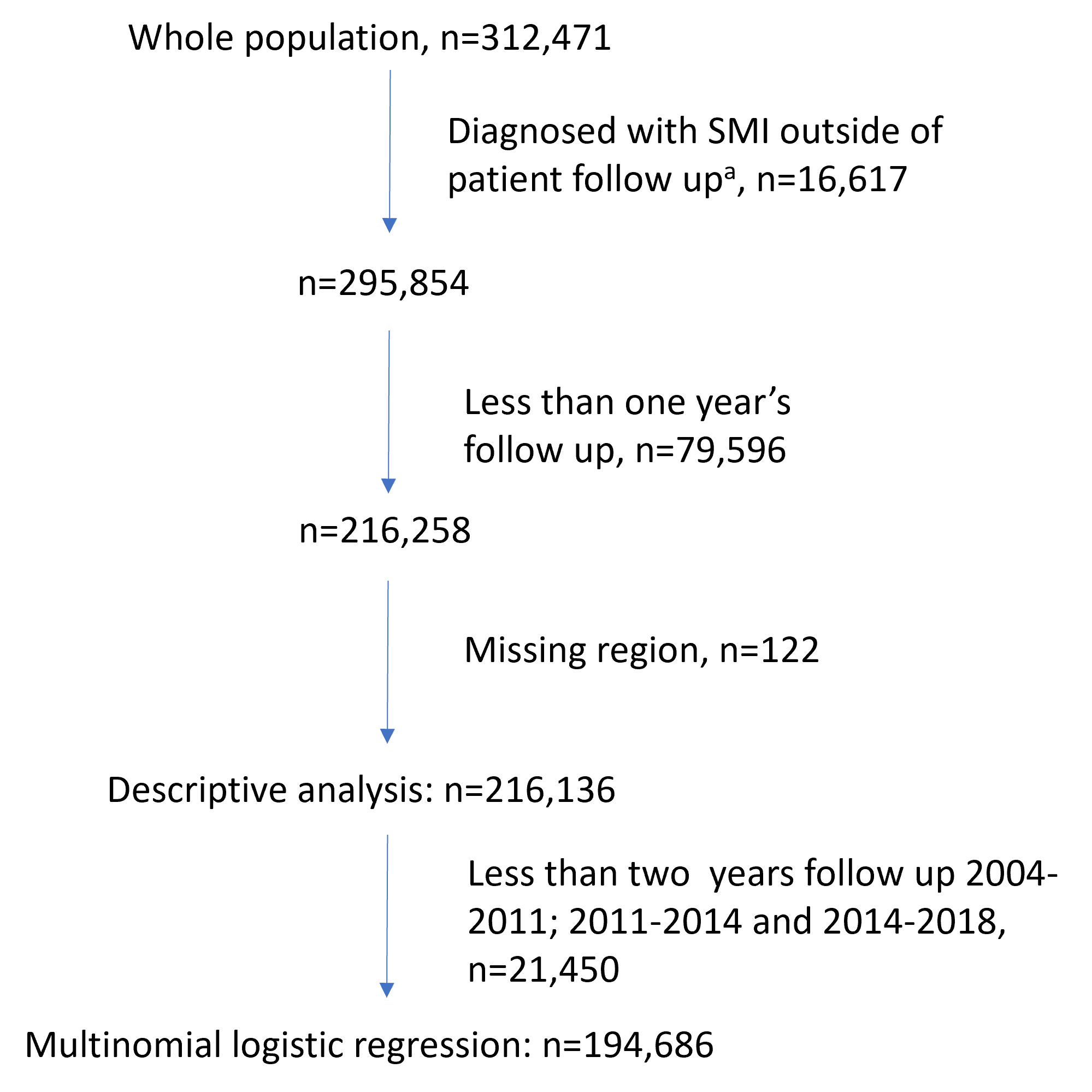

a: After follow-up ends (earliest of death, leaving the primary care practice, age 100 or last data collection by CPRD) or under 18.

Figure S2: Cardiovascular risk factor screening prevalence in patients with severe mental illness, by age

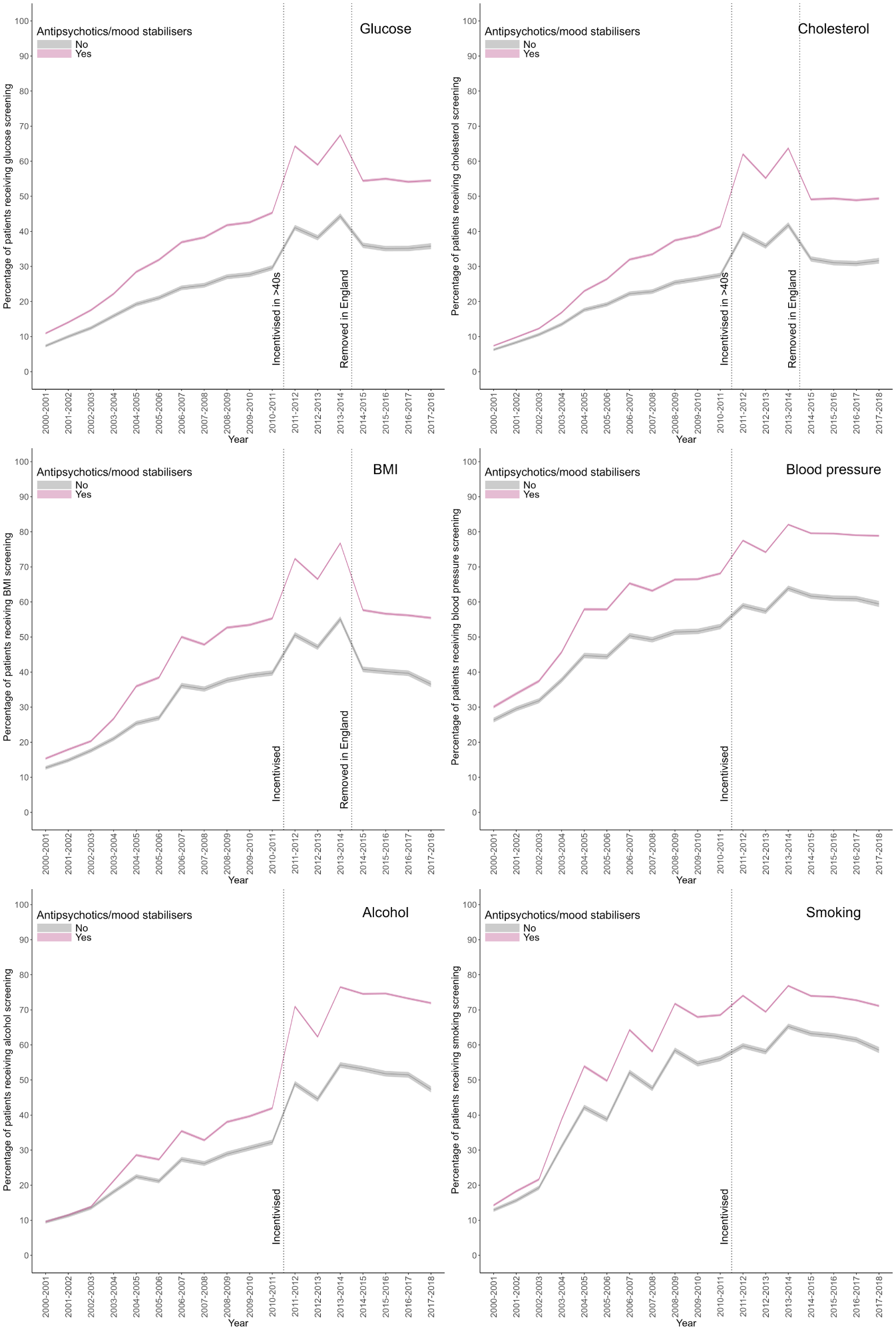

Figure S3: Cardiovascular risk factor screening prevalence in people with severe mental illness, by country*

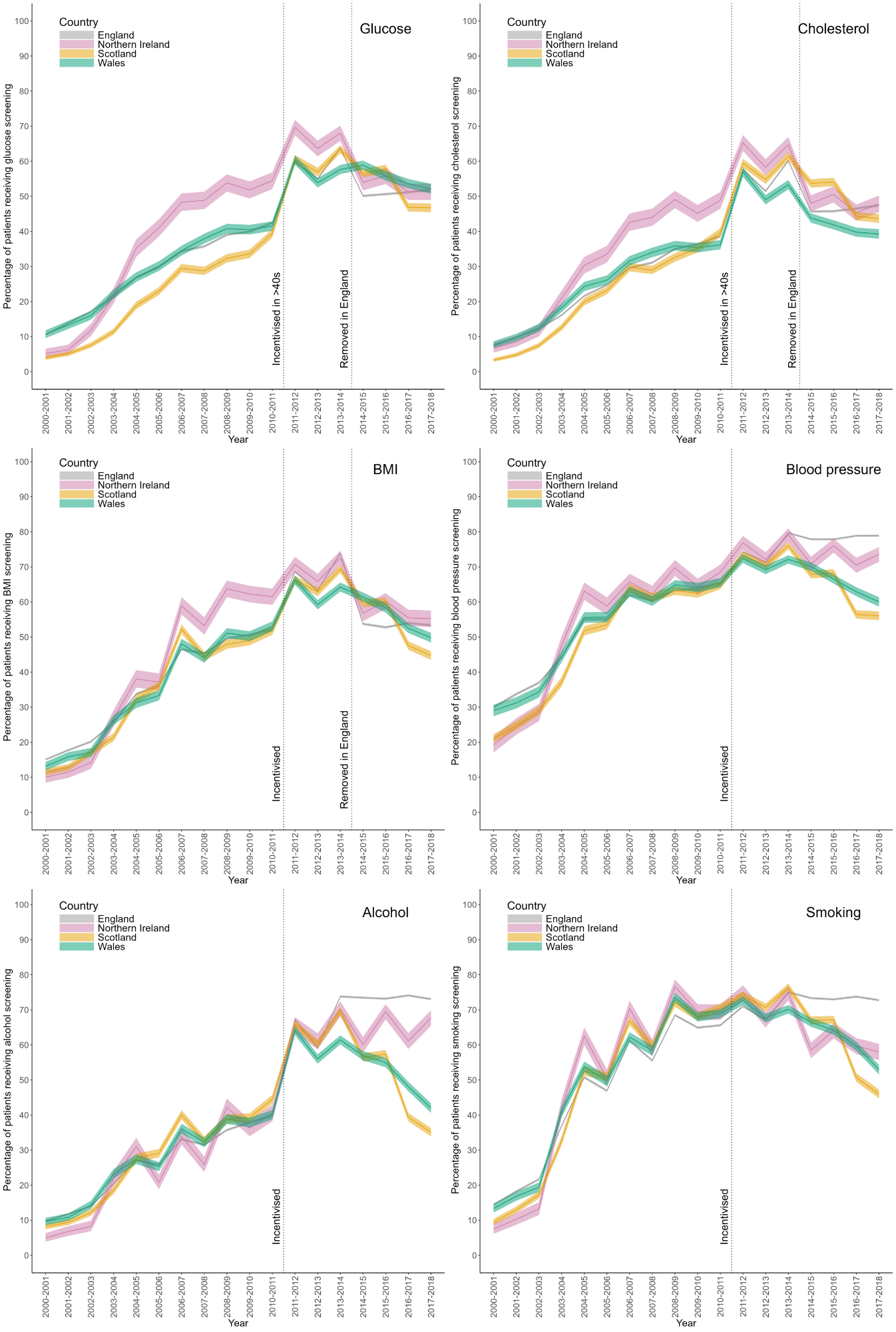

*From 2011-2012 incentivisation differs by country. See Table S1 for further explanation

Figure S4: Cardiovascular risk factor screening prevalence in people with severe mental illness, by current severe mental illness diagnosis

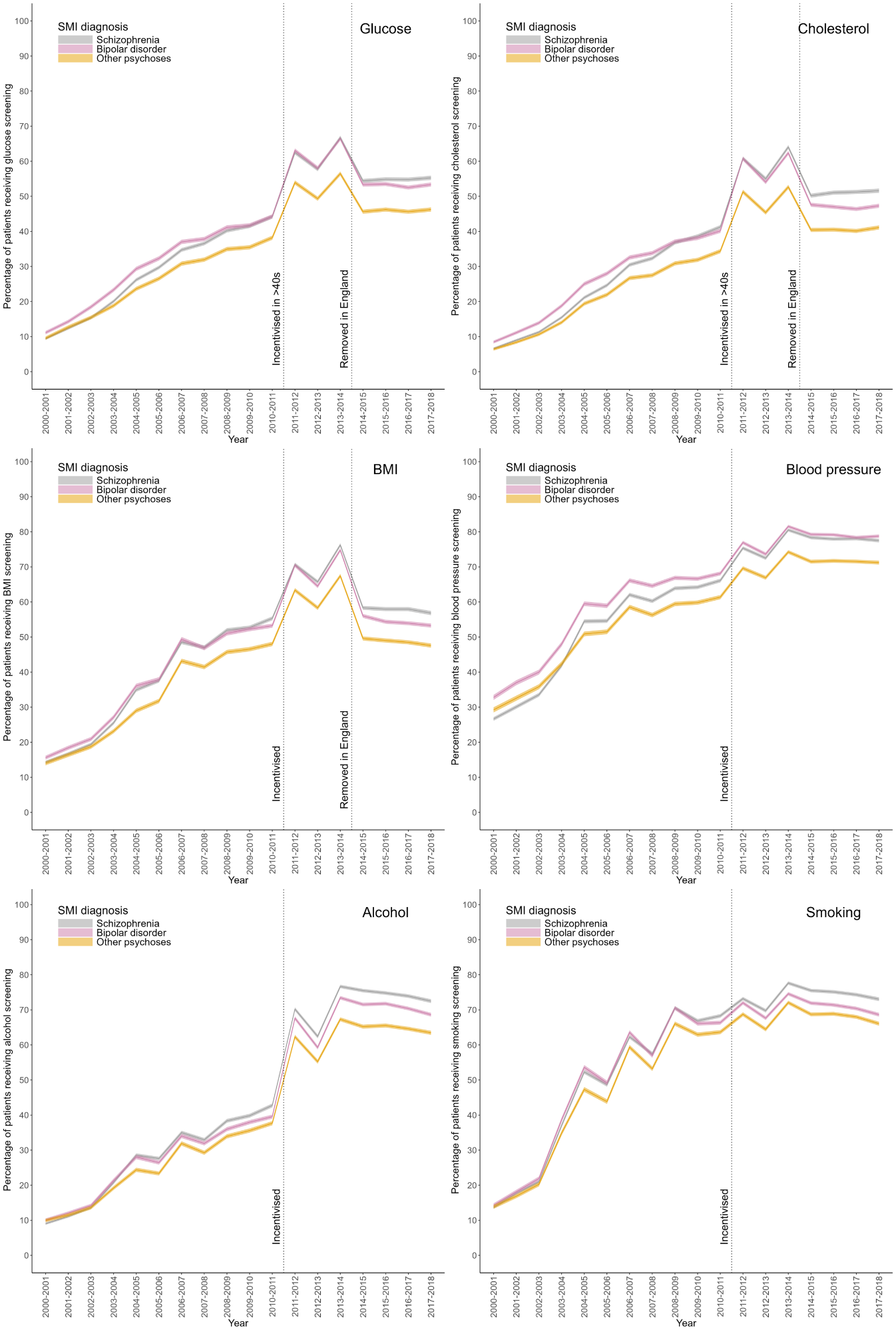

Figure S5: Cardiovascular risk factor screening prevalence in people with severe mental illness, by sex

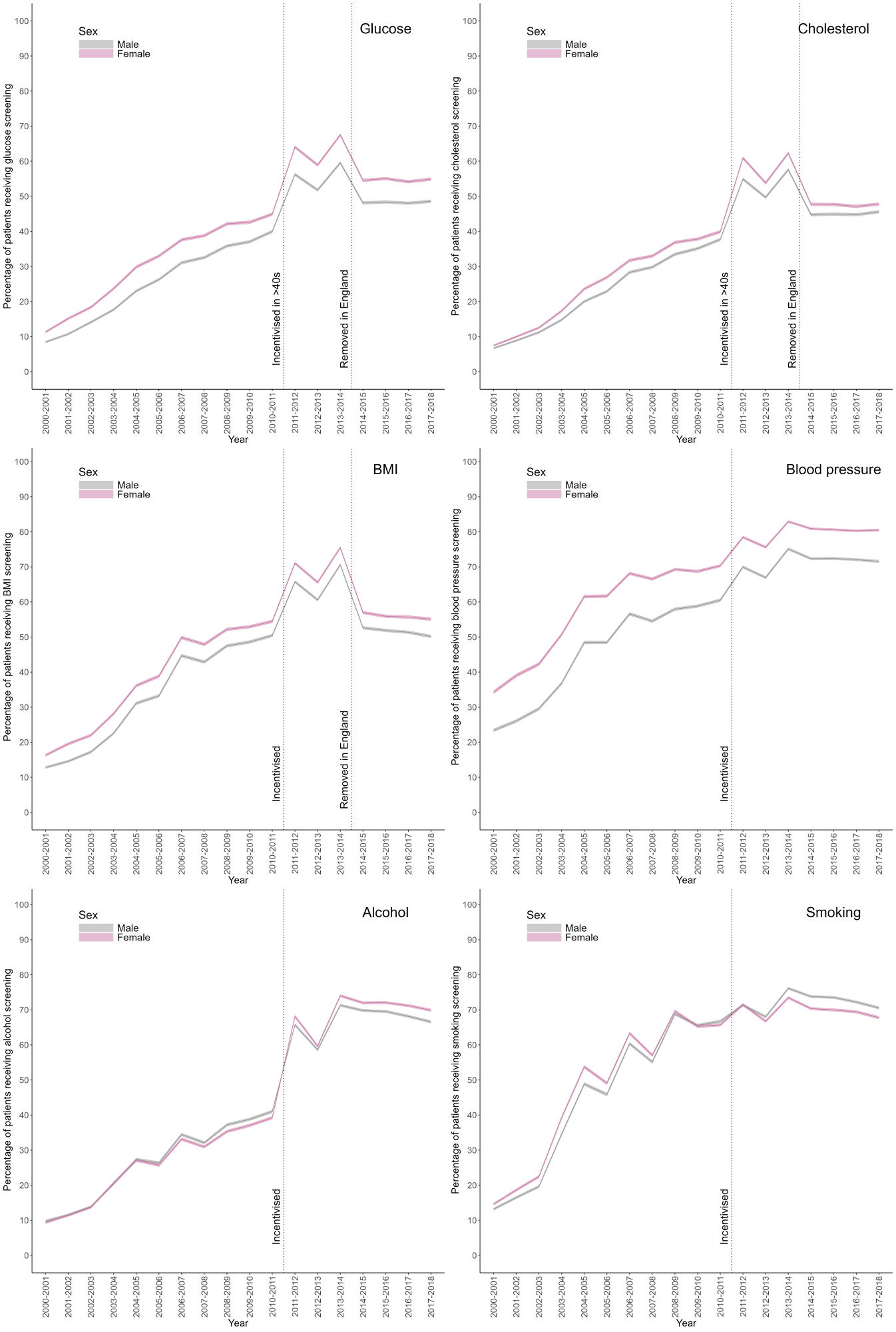

Figure S6: Cardiovascular risk factor screening prevalence in people with severe mental illness, by presence on another QOF register that incentivises cardiovascular risk factor screening

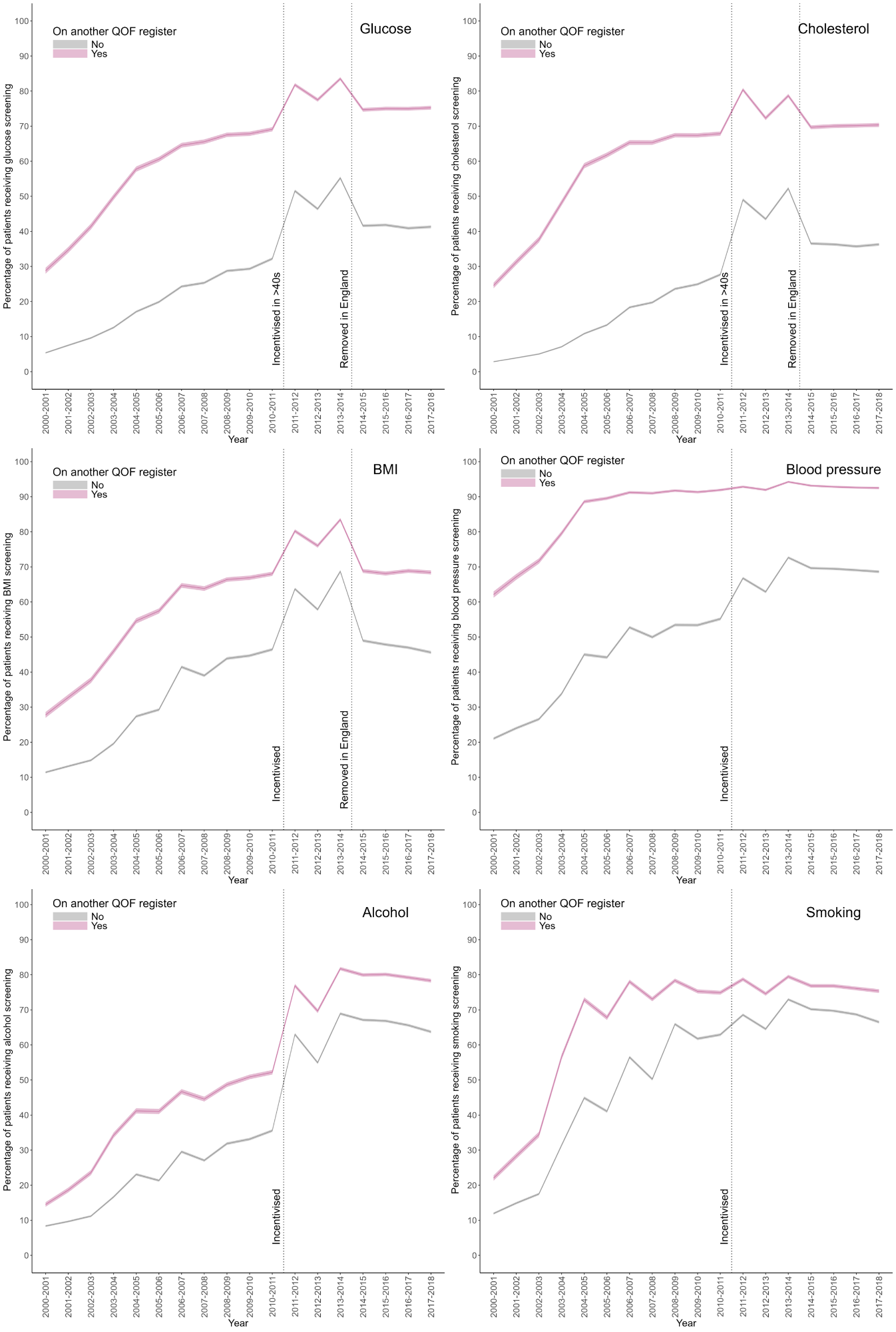

Figure S7: Cardiovascular risk factor screening prevalence in people with severe mental illness, by prescription of antipsychotics or mood stabilisers

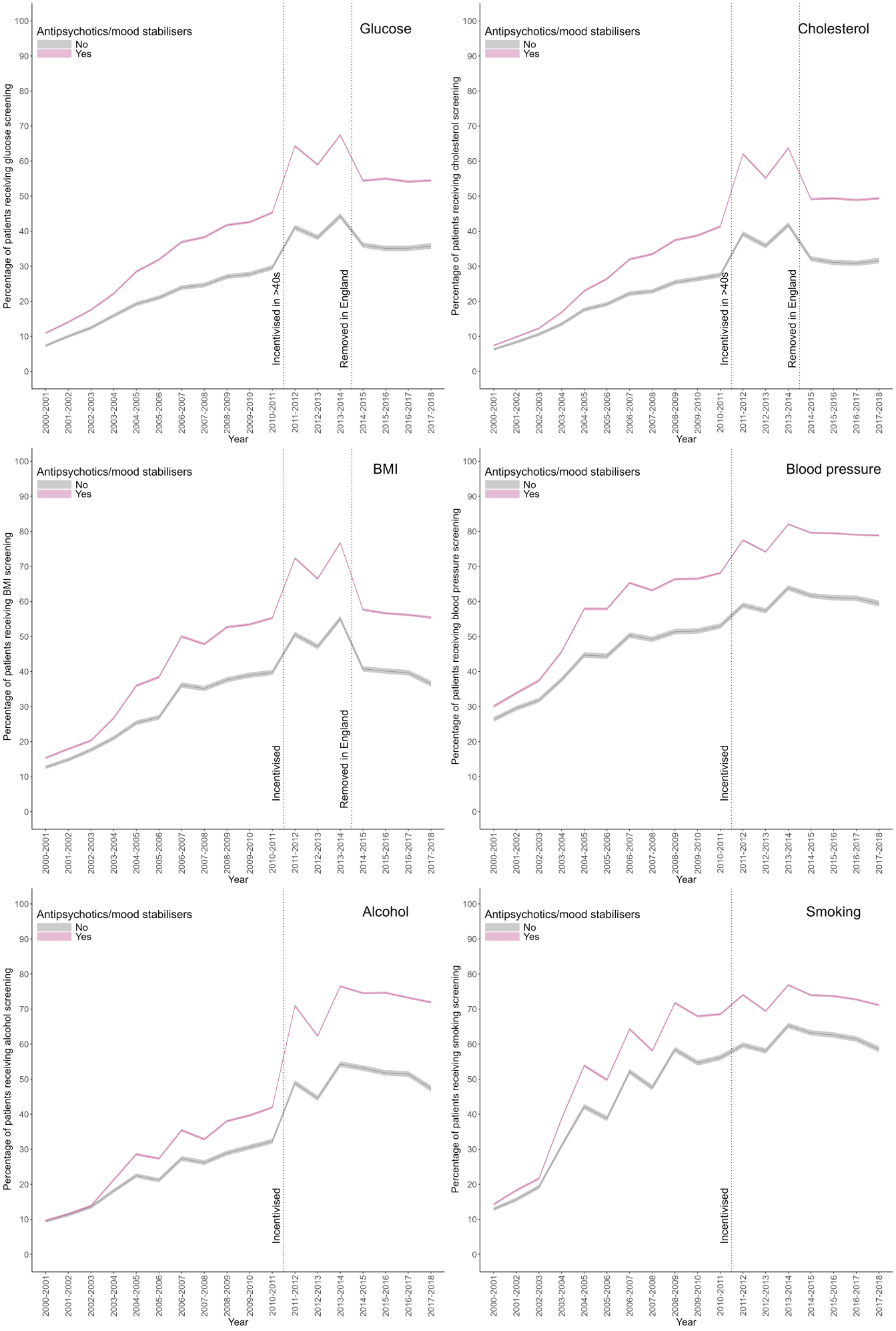

Figure S8: Cardiovascular risk factor screening in patients with severe mental illness, by ethnicity

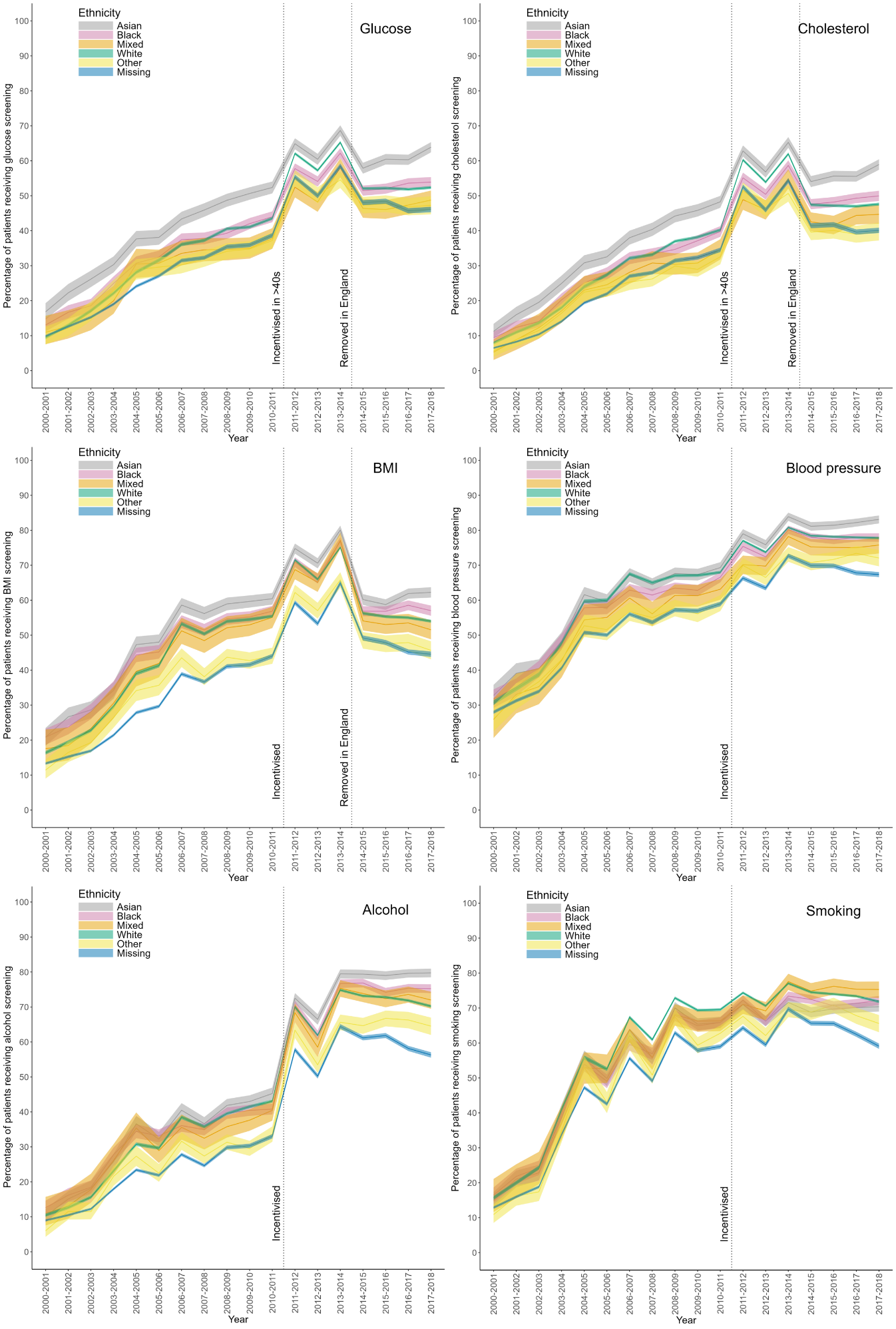

Figure S9: Cardiovascular risk factor screening prevalence in patients with severe mental illness, by exception reporting status that year*

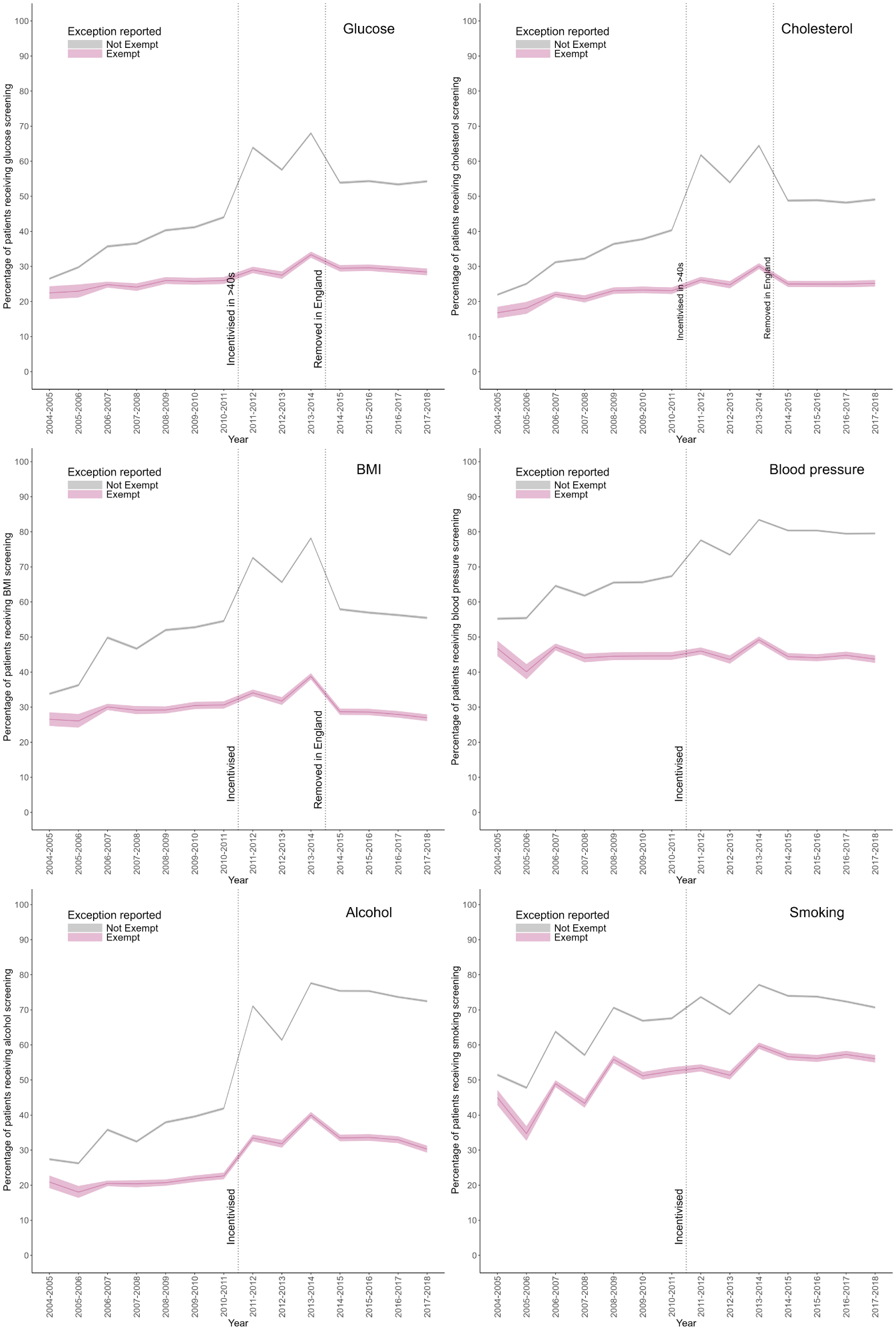

*Limited to 2004 onwards when exception reporting was introduced

Figure S10: Cardiovascular risk factor screening prevalence in people with severe mental illness, by index of multiple deprivation for patients in England where this is available (n=137,941)*
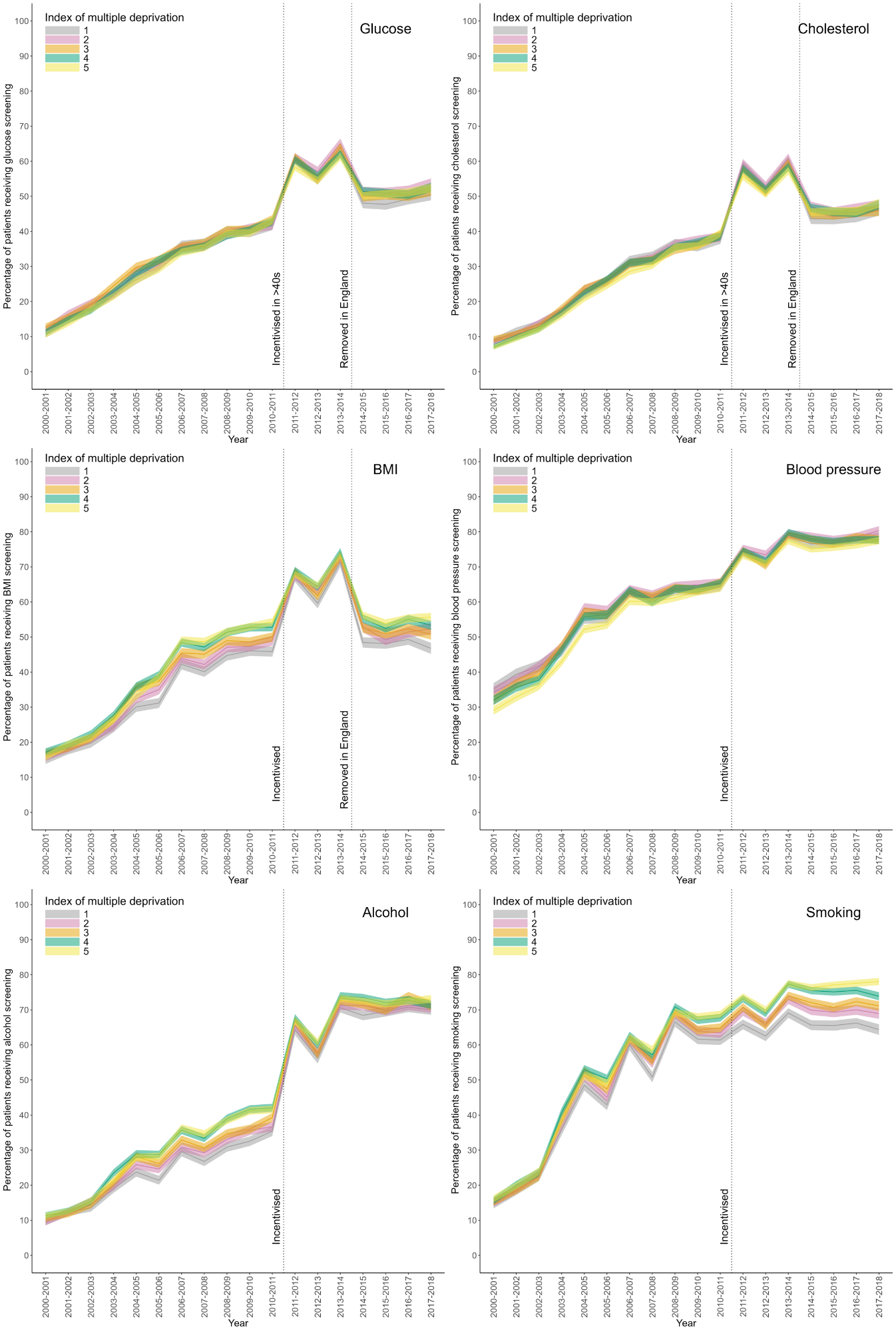
 * 1=least deprived; 5=most deprived.
